# Climatic and socio-economic factors supporting the co-circulation of dengue, Zika and chikungunya in three different ecosystems in Colombia

**DOI:** 10.1101/2020.11.18.20233940

**Authors:** Jasmine Morgan, Clare Strode, J. Enrique Salcedo-Sora

**Author notes:** Corresponding authors (CS).

## Abstract

Dengue, Zika and chikungunya are diseases of global health significance caused by arboviruses and transmitted by the mosquito *Aedes aegypti* of worldwide circulation. The arrival of the Zika and chikungunya viruses to South America increased the complexity of transmission and morbidity caused by these viruses co-circulating in the same vector mosquito species. Here we present an integrated analysis of the reported arbovirus cases between 2007 and 2017 and local climate and socio-economic profiles of three distinct Colombian municipalities (Bello, Cúcuta and Moniquirá). These locations were confirmed as three different ecosystems given their contrasted geographic, climatic and socio-economic profiles. Correlational analyses were conducted with both generalised linear models and generalised additive models for the geographical data. Average temperature and wind speed were strongly correlated with disease incidence. The transmission of Zika during the 2016 epidemic appeared to decrease circulation of dengue in Cúcuta, an area of sustained high incidence of dengue. Socio-economic factors such as barriers to health and childhood services, inadequate sanitation and poor water supply suggested an unfavourable impact on the transmission of dengue, Zika and chikungunya in all three ecosystems. Socio-demographic influencers were also discussed including the influx of people to Cúcuta, fleeing political and economic instability from neighbouring Venezuela. *Aedes aegypti* is expanding its range and increasing the global threat of these diseases. It is therefore vital that we learn from the epidemiology of these arboviruses and translate it into an actionable knowledge base. This is even more acute given the recent historical high of dengue cases in the Americas in 2019, preceding the COVID-19 pandemic, which is itself hampering mosquito control efforts.

**Author summary:** Viruses transmitted by *Ae. aegypti* mosquitoes (dengue, Zika, chikungunya) are amongst the most significant public health concerns of recent years due to the increase in global cases and the rapid spread of the vectors. The primary method of controlling the spread of these arboviruses is through mosquito control. Understanding factors associated with risk of these viruses is key for informing control programmes and predicting when outbreaks may occur. Climate is an important driver in mosquito development and virus reproduction and hence the association of climate with disease risk. Socio-economic factors contribute to perpetuate disease risk. Areas of high poverty have abundance of suitable habitat for *Ae. aegypti* (e.g. due to poor housing and sanitation). This study investigated the factors effecting arbovirus incidence in three distinct regions of Colombia: Bello, Cúcuta and Moniquirá. The results show significant relationships between disease incidence and temperature, precipitation and wind speed. A decline in dengue following outbreaks of Zika (2016) is also reported. Measures of poverty, including critical overcrowding and no access to improved water source were also found to be higher in areas of higher disease incidence. The results of this study highlight the importance of using a multifactorial approach when designing vector control programs in order to effectively distribute health care resources.

## Introduction

Vector-borne diseases are one of the most significant public health burdens globally, with 80% of the total world population at risk [1]. Arboviruses, including dengue, Zika and chikungunya, are of particular concern due to the recent increase in global cases promoted by the rapid spread of both, their primary mosquito vector *Aedes aegypti* as well as their secondary vector *Aedes albopictus* [2]. Dengue infection can be asymptomatic but clinical presentations range from mild dengue fever (DF), a febrile illness similar to influenza, to the severe forms of dengue; dengue shock syndrome (DSS) and dengue haemorrhagic fever (DHF) [3]. Most Zika (ZIKV) infections are asymptomatic, with only approximately 20% of infections causing symptoms [4,5]. The clinical presentations of symptomatic ZIKV can include Zika fever, congenital Zika syndrome and Guillain-Barré syndrome. Congenital Zika syndrome refers to a group of birth defects, notably Microcephaly, which have been associated with ZIKV during pregnancy [6,7]. Infection with the chikungunya virus (CHIKV) is characterised by sudden onset fever, rash and arthralgia [8].

Dengue causes an estimated 390 million infections per year and has a distribution that covers every continent of the world with the exception of Antarctica [9]. The number of global dengue cases reported to the WHO has increased 15 fold over the last 20 years, with deaths also seeing a significant increase (4-fold) [10]. The first epidemics of ZIKV were reported in Yap, Micronesia (2007) and French Polynesia (2013), outbreaks were reported in Brazil in 2015 and 2016 which then led to a rapid spread of ZIKV to 48 countries within the Americas and the Caribbean [11]. ZIKV epidemics have also been reported in Singapore [12], Vietnam [13], Thailand [14] and Cape Verde [15]. CHIKV was first reported in Tanzania in 1952 and has since rapidly spread across the globe causing epidemics in Asia, India, Europe and The Americas [16].

Spreading from its ancestral home in West Africa 400-500 years ago via the slave trade *Ae. aegypti* is found in tropical and sub-tropical regions [17]. Meteorological conditions directly influence the incidence of arboviruses by modulating vector mosquito populations. Conditions favourable to *Ae. aegypti* include an ideal temperature range between 20 - 35°C for mosquito development, fertilisation and viral load [18]. Non-climatic factors promoting *Ae. aegypti* populations include vegetation index, urbanisation and accessibility to human populations [19,20]. Latin America is significantly affected by *Ae. aegypti* borne viruses due to its habitat suitability for the vector, tropical climate and often limited medical resources and vector control programmes [21,22]. Colombia, located in the north-west corner of South America, has 140,612 km^2^ of suitable habitat for *Ae. aegypti* throughout the country based on the presence of climatic characteristics [23]. *Ae. aegypti* in Colombia has been found at altitudes up to 2,300 m above sea level [24]. The presence of *Ae. aegypti* across Colombia is mirrored by a high nationwide incidence of dengue, Zika and chikungunya. Dengue has been consistently reported in Colombia over the past two decades causing an average of 84,926 cases each year (1980-2019). Zika was first reported in Colombia in 2015 and was followed by a significant outbreak of 91,711 cases in 2016. Chikungunya was first detected in Colombia in 2013, causing 275,907 cases in that single year [25].

In addition to climatic variables, socio-economic (SE) factors can contribute to the spread of mosquito borne diseases. This is particularly acute with *Ae. aegypti*, a highly anthropophilic species which lives within or in close proximity to human dwellings breeding in domestic water storage containers. Poor housing construction together with high population density and inadequate sanitation with little to no access to clean running water are key SE factors promoting *Ae. aegypti* populations [26]. In the absence of suitable vaccines for dengue, Zika or chikungunya, disease prevention is currently based on *Ae. aegypti* control. This is challenging with a diurnal biting mosquito. A compound effect is the development of insecticide resistance in populations of *Ae. aegypti* reported in areas of Colombia [27].

This study aims to investigate the epidemiology of these three arboviruses (dengue, Zika and chikungunya) co-circulating in a single vector species (*Ae. aegypti*) in three distinct eco-systems in Colombia between 2007 - 2017. This is achieved using a multifactorial approach considering the potential correlations of several meteorological and socio-economic factors with disease incidence. We show that specific climatological factors are strong drivers for these arboviral diseases to which contextual socio-economical characteristics can act as modifiers. Importantly, we find a discriminatory pattern between these three diseases highlighting unexpected dynamics of transmission between Zika and dengue particularly in an area of high dengue circulation.

## Methods

### Disease incidence data

Three municipalities in Colombia were selected for analysis: Bello, Cúcuta and Moniquirá (Fig 1). Cases of dengue, severe dengue (dengue shock syndrome and dengue haemorrhagic fever), chikungunya and Zika reported in each epidemiological week were obtained for the period of 2007 to 2017 from SIVIGILIA (National Public Health Surveillance System, Colombian National Institute of Health) [28]. This period of study (2007-2017) had consistent reporting of dengue along with the epidemics of both Zika and chikungunya. Chikungunya was first reported in 2014 and Zika in 2015. For the purpose of analysis week 53 was removed from any years in which there were 53 epidemiological weeks (2008 and 2014) this ensured continuity across the data set with all years comprising of 52 weeks when analysed. Only cases confirmed by SIVIGILIA were used in this analysis, confirmations are made based on laboratory tests and epidemiological links.

**Fig 1.**
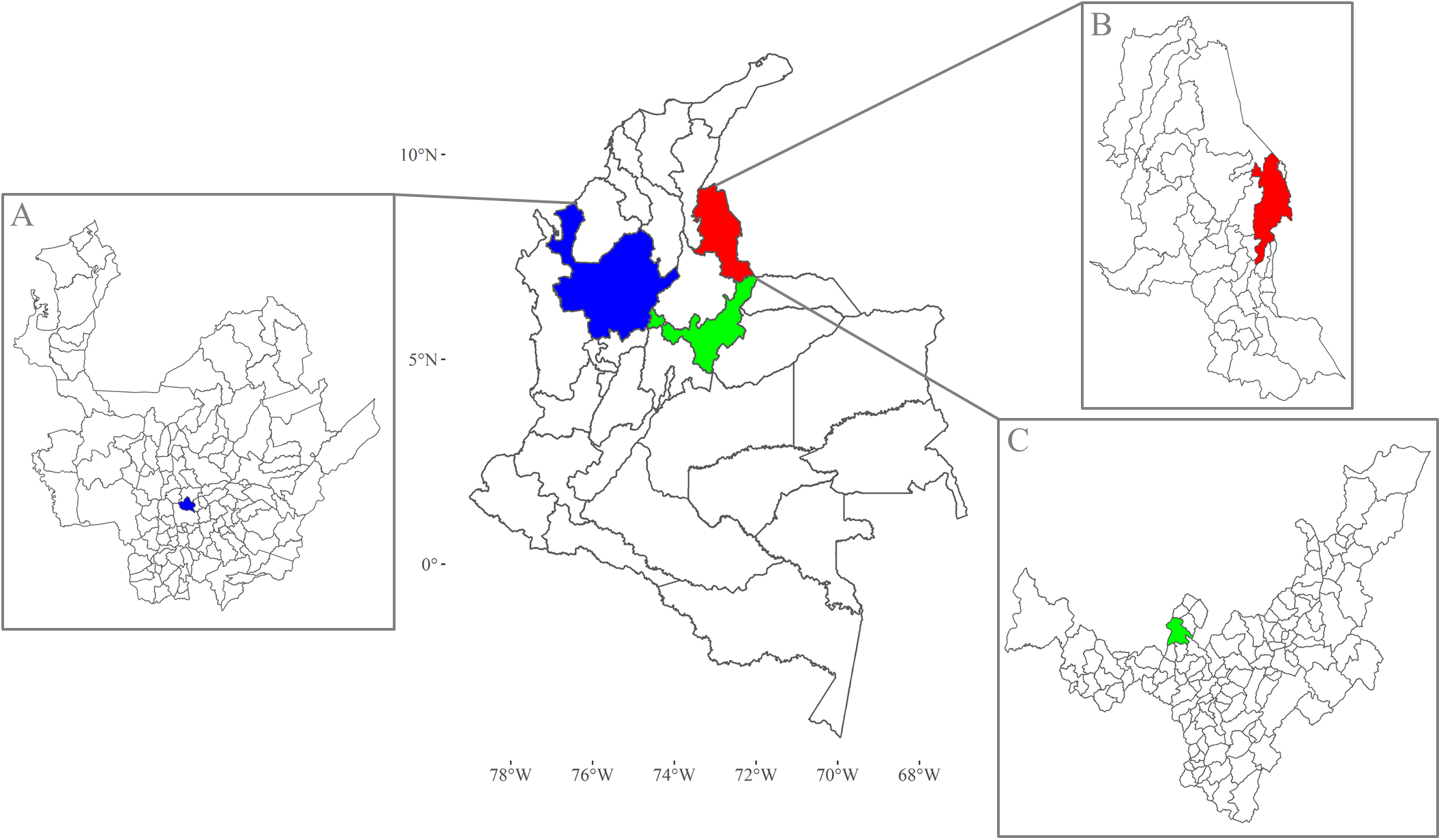
Map of Colombia showing the location of each municipality within their political divisions (departments). Departments are the largest units of local government answerable to the country’s national government. (A) Department of Antioquia governs Bello which is denoted as a small blue area. (B) Department of Norte de Santander has as its capital Cúcuta, a city (red) to the East of this department that shares the border with Venezuela. (C) Department of Boyacá has the municipality of Moniquirá (green).

### Climate data

Daily weather data for Bello, Cúcuta and Moniquirá were obtained from the NASA Langley Research Center (LaRC) POWER Project [29]. The meteorological data has a resolution of 0.5° latitude and 0.5° longitude. The weather data was obtained for the same time period as the disease incidence data (2007-2017) and converted to epidemiological weeks to correspond with the dates of the disease incidence data. The weather variables included for each municipality were: maximum temperature (Tmax), minimum temperature (Tmin), average temperature (Tavg), maximum wind speed (WSmax), minimum wind speed (WSmin), average wind speed (WSavg), total precipitation and average humidity (Havg).

### Population and socio-economic data

Population data for each municipality were obtained using population projections by Colombia’s National Administrative Department of Statistics (Departamento Administrativo Nacional de Estadística) (DANE) (www.dane.gov.co) [30]. The multidimensional poverty index (MPI) was implemented by the Oxford Poverty and Human Development Initiative and the United Nations Development Program’s Human Development Report Office as a direct method for measuring poverty [31]. The MPI at municipality level was obtained from DANE using data collected in the 2018 National Population and Housing Census and using the indicators and respective weightings listed in S2 Table [32].

### Statistical analysis

#### Patterns of disease incidence by location

Differences in the total burden of all three *Ae. aegypti* borne viruses as well as the individual burden of dengue and severe dengue were investigated using the total number of cases from 2007-2017. Poisson Generalised Linear Models (GLMs) were initially carried out, revealing overdispersion (data variance greater than expected for the given model) statistics of 545, 542 and 123 for total disease, dengue and severe dengue respectively. To correct for the large overdispersion the GLMs were recalculated with negative binomial errors [33] using the glm.nb function of the R package MASS [34]. Differences between all three municipalities were tested with Tukey pairwise comparisons using the glht function of the multcomp R package [35]. Incidence of chikungunya in 2015 was initially modelled using a Poisson GLM which revealed an overdispersion statistic of 2.9. The standard errors were therefore corrected using quasi-GLMs where the variance was theta x mu. Where mu was the mean of the dependant variable and theta the dispersion parameter of the quasi-GLMs [33]. Quasi-GLMs were conducted using the glm function from the R package stats [36]. Zika incidence was modelled for the year 2017 only, initially a Poisson GLM was used and an overdispersion statistic of 99 was detected. As the overdispersion statistic was above 20 it was corrected for using negative binomial errors [33]. Total *Ae. aegypti* borne disease and dengue incidence were also modelled for 2015 and 2016 using quasi-GLMs to account for low level overdispersion except for total disease in 2016 which had a dispersion statistic of 90 and was therefore modelled with a negative binomial GLM. Population was used as an offset in all models in order to standardise disease incidence by population size.

#### Patterns of disease incidence over time

For the pattern of disease over time we used total yearly incidence data. Poisson GLMs were initially used to model each disease in each municipality but overdispersion was detected in some models, hence error distributions were adjusted accordingly. For Bello incidence of both total disease (dengue, severe dengue, chikungunya and Zika) and dengue alone were modelled using quasipoisson GLMs, correcting for overdispersions of 3.69 and 3.63 respectively. For Cúcuta a negative binomial GLM was required for total disease incidence in order to correct for overdispersion of 32.52 and quasi-GLM was used for dengue incidence due to slight overdispersion of 4.87. Severe dengue incidence in Cúcuta was not found to be significantly overdispersed when modelled with a Poisson GLM (overdispersion statistic = 1.79), as the overdispersion statistic was <2. For Moniquirá both total disease incidence and dengue incidence alone were modelled using quasi-GLMs, correcting for respective overdispersion statistics of 2.39 and 2.34 [33]. Population was used as an offset in all models in order to standardise disease incidence by population size. All quasi-GLMs were conducted using the glm function from the R package stats [36] and negative binomial GLMs used the glm.nb function of the R package MASS [34]. Differences between the years were tested with Tukey pairwise comparisons using the glht function of the multcomp R package [35].

#### Generalised additive models

The correlations between climatic variables and the total disease incidence (dengue, severe dengue, chikungunya and Zika) across all three locations were investigated using a generalised additive model (GAM). The weekly disease incidence and weather data for each municipality was converted into 4-week data, matching the dates of epidemiological months. Combining the data into 4-week periods rather than individual weeks reduced zero inflation improving the reliability of the GAM outputs. All climate variables were lagged by plausible time lags for their effect on disease incidence, of 4 and 8 weeks. Square root transformations were used for total disease incidence and each weather variable due to non-normal distribution. Generalised additive models were chosen due to their ability to model non-linear relationships between a response variable (disease incidence) and multiple explanatory variables (climate variables) [37]. A quasi-maximum likelihood Poisson GAM was used in order to prevent possible overdispersion [33]. Population size was used as an offset to standardise disease incidence by population. Initially all climate variables with both 4 and 8-week time lags were assumed to have a non-linear relationship and were therefore modelled as smoothed terms. Subsequent analysis of the effective degrees of freedom (*edf*) was used to identify variables with *edf* = 1.0, suggesting linearity. These variables were then included in the model as linear rather than smoothed terms. Generalised cross validation (GCV) was used to determine the most appropriate model. Generalised additive modelling and subsequent model validation was conducted using the R package mgcv [38]. Visualisation of GAM estimations were conducted using the mgcViz R package [39].

#### Socio-economic factors

Principle components analysis (PCA) was used for dimensional reduction to allow the inclusion of socio-economic data with previously compiled geographic and climate data. The PCA was conducted using the R package stats [36] and visualisations of the PCA were created using the factoextra R package [40].

## Results

### Study Locations

The three municipalities Bello, Cúcuta and Moniquirá were chosen due to their geographical separation, and distinct climate characteristics, demographics and burden of *Ae. aegypti* borne diseases (Table 1).

**Table 1.** Climate, population and disease incidence for each municipality. Elevation, yearly precipitation, mean temperature and humidity were calculated from climatological data for 1981-2010 [41]. Population for each municipality calculated from 1985-2020 population projections (Source: National Administrative Department of Statistics: www.dane.gov.co [30]). Disease incidence: Burden of *Ae. aegypti* borne diseases in cases per 100,000 people [28].

| Municipality | Elevation (m) | Annual Precipitation (mm) | Mean Temperature (°C) | Mean Relative Humidity (%) | Climate Classification (Caldas-Lang) | Population | Disease Incidence |
| --- | --- | --- | --- | --- | --- | --- | --- |
| Bello | 1438 | 1542.44 | 21.96 | 76.80 | Temperate Semi-Wet | 356,504 | 1301 |
| Cúcuta | 250 | 904.18 | 27.19 | 71.05 | Warm Semiarid | 558,599 | 6093 |
| Moniquirá | 1700 | 2006.25 | 19.09 | 76.53 | Cold Humid | 21,249 | 4669 |

### General disease incidence between 2007 - 2017

Dengue was the most prevalent of the three diseases throughout Colombia (Fig 2). Cúcuta carried the highest disease burden of the three municipalities followed by Moniquirá, with Bello having the lowest disease incidence. The total number of confirmed cases of all *Ae. aegypti* borne diseases (dengue, severe dengue, chikungunya and Zika) during the period of 2007 - 2017 were 5,727, 32,328 and 1,005 in Bello, Cúcuta and Moniquirá respectively. The breakdown of cases per 100,000 people of all three diseases between 2007 and 2017 in these locations were 1,301, 6,094 and 4,669 in Bello, Cúcuta and Moniquirá, respectively. When discriminating by disease per 100,000 people, the incidence of dengue was 1,263 in Bello, 5,106 in Cúcuta and 4,566 in Moniquirá. Chikungunya had 27 cases in Bello, 154 in Cúcuta and 61 Moniquirá. Incidence of Zika was lowest in Bello with 11 cases compared to 834 in Cúcuta and 42 in Moniquirá.

**Fig 2:**
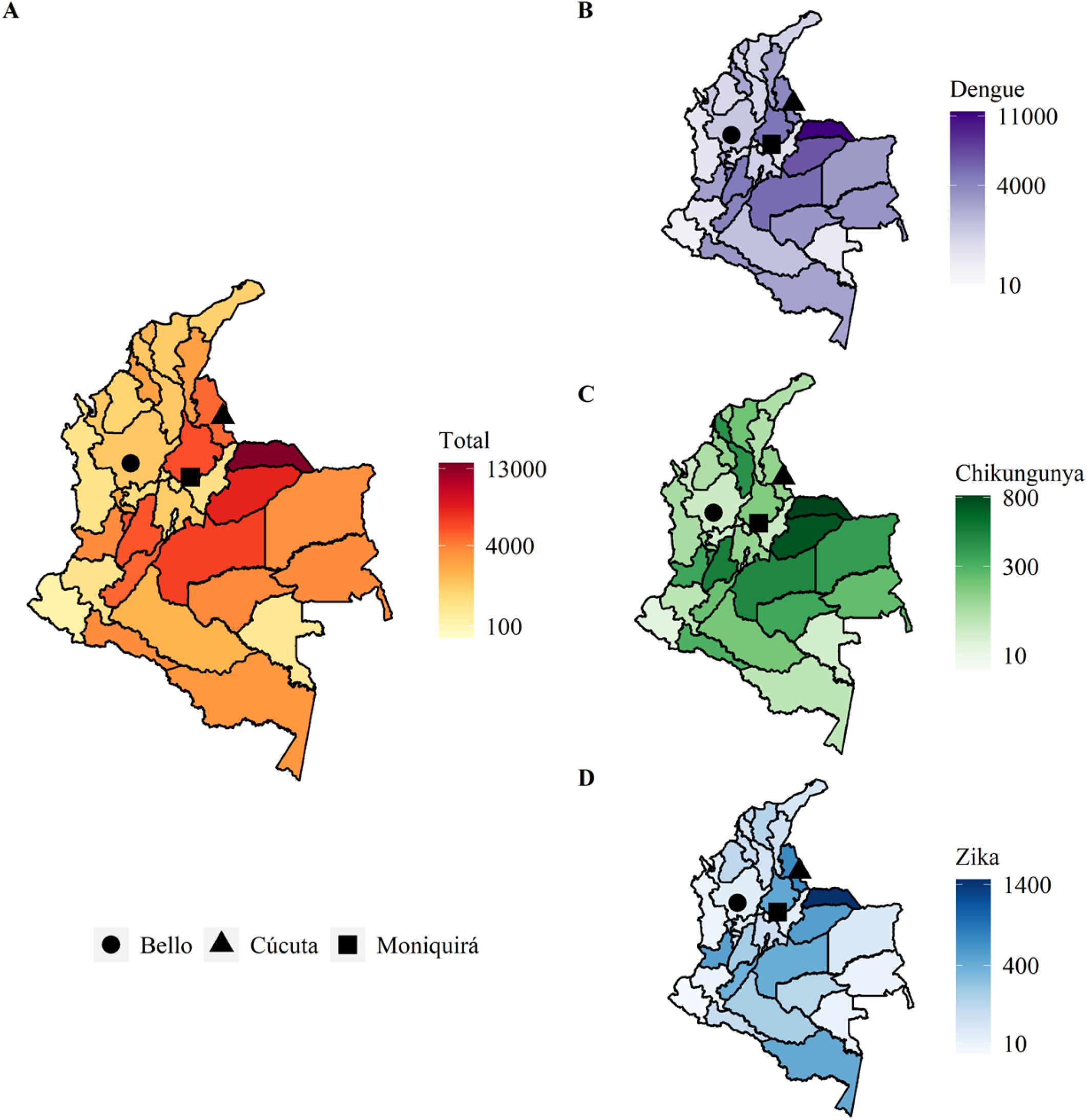
Arboviral diseases in Colombia per 100,000 population for the period of 2007-2017. (A) Number of total *Ae. aegypti* borne diseases in Colombia. (B) Number of cases of dengue, (C) chikungunya and (D) Zika.

The spread of the data for the period analysed was assessed in 12-week periods, using smooth moving averages (SMA) (Fig 3). Dengue and severe dengue were consistently reported throughout the period of 2007-2017. Dengue data showed at least three spikes: one in 2009 (approximately week 180 in Fig 3), followed by two more by the end of 2014 (week 400) and beginning of 2017 (week 510). The latter of these peaks started approximately two years prior (2015) (Fig 3). Chikungunya cases were only reported between 2014-2017, hence the flat line before week 400 in Fig 3. Similarly, for Zika whose first cases were reported in 2015 (approximately week 450 in Fig 3), with the highest number of cases reported in week 468 in 2016 (Fig 3).

**Fig 3:**
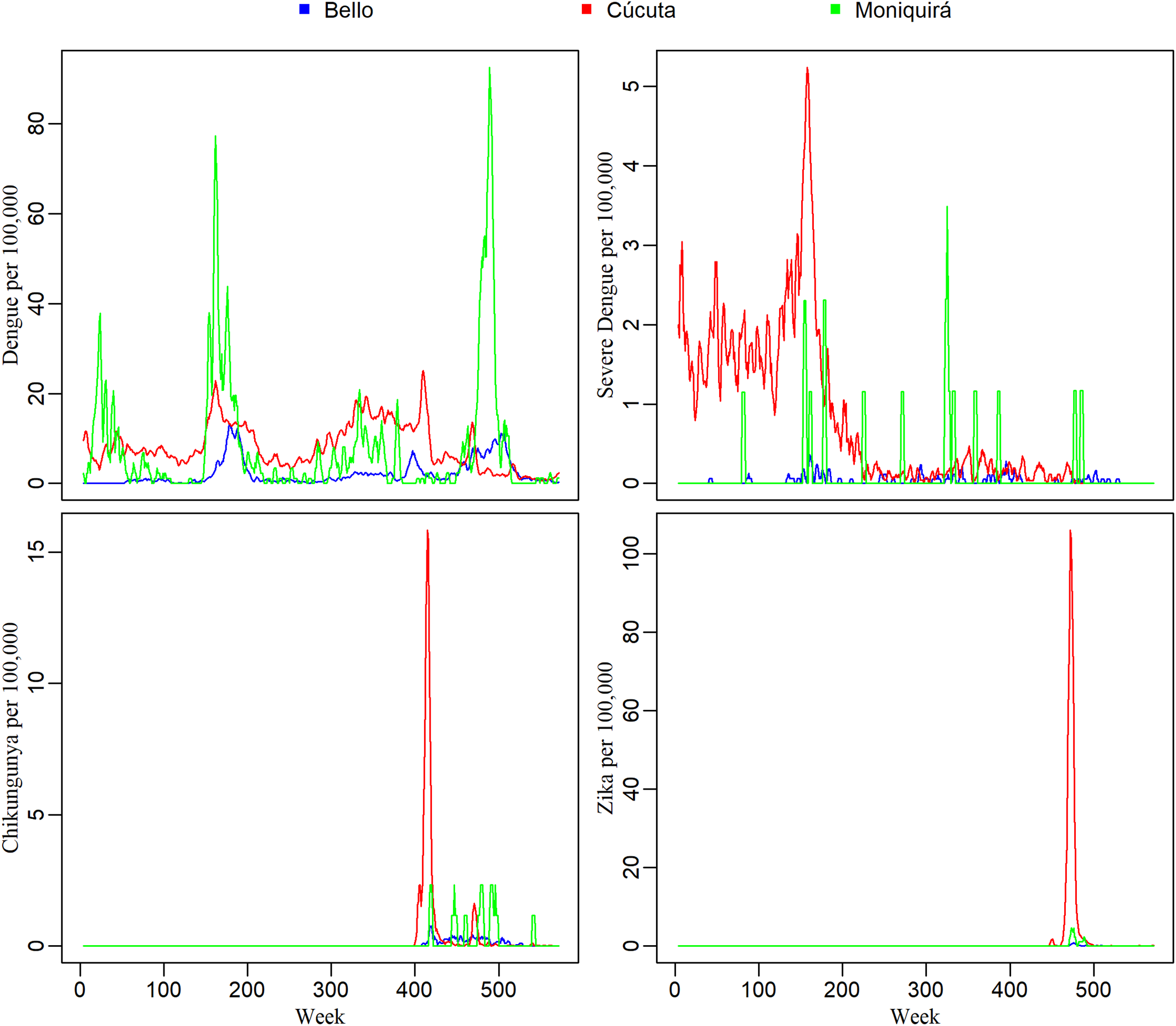
Data granularity for disease reported from 2007 -2017. Smooth moving averages (SMA) 12-week time series of dengue, severe dengue, chikungunya and Zika cases per 100,000 people in Bello, Cúcuta and Moniquirá.

### Patterns of disease incidence per location

Total disease incidence over the 11-year period was significantly lower in Bello than in Cúcuta (p = < 0.001) and Moniquirá (p = 0.005) (Fig 4A) for all three diseases. The number of dengue cases were similarly high between Moniquirá and Cúcuta (*p* = 0.99). However, severe dengue incidence was significantly different across all three municipalities: Cúcuta had the highest burden of severe dengue when compared to both Bello (p = <0.001) and Moniquirá (p = 0.005), and Moniquirá had a significantly higher burden of severe dengue when compared to Bello (p = 0.047) (Fig 4A).

**Fig 4:**
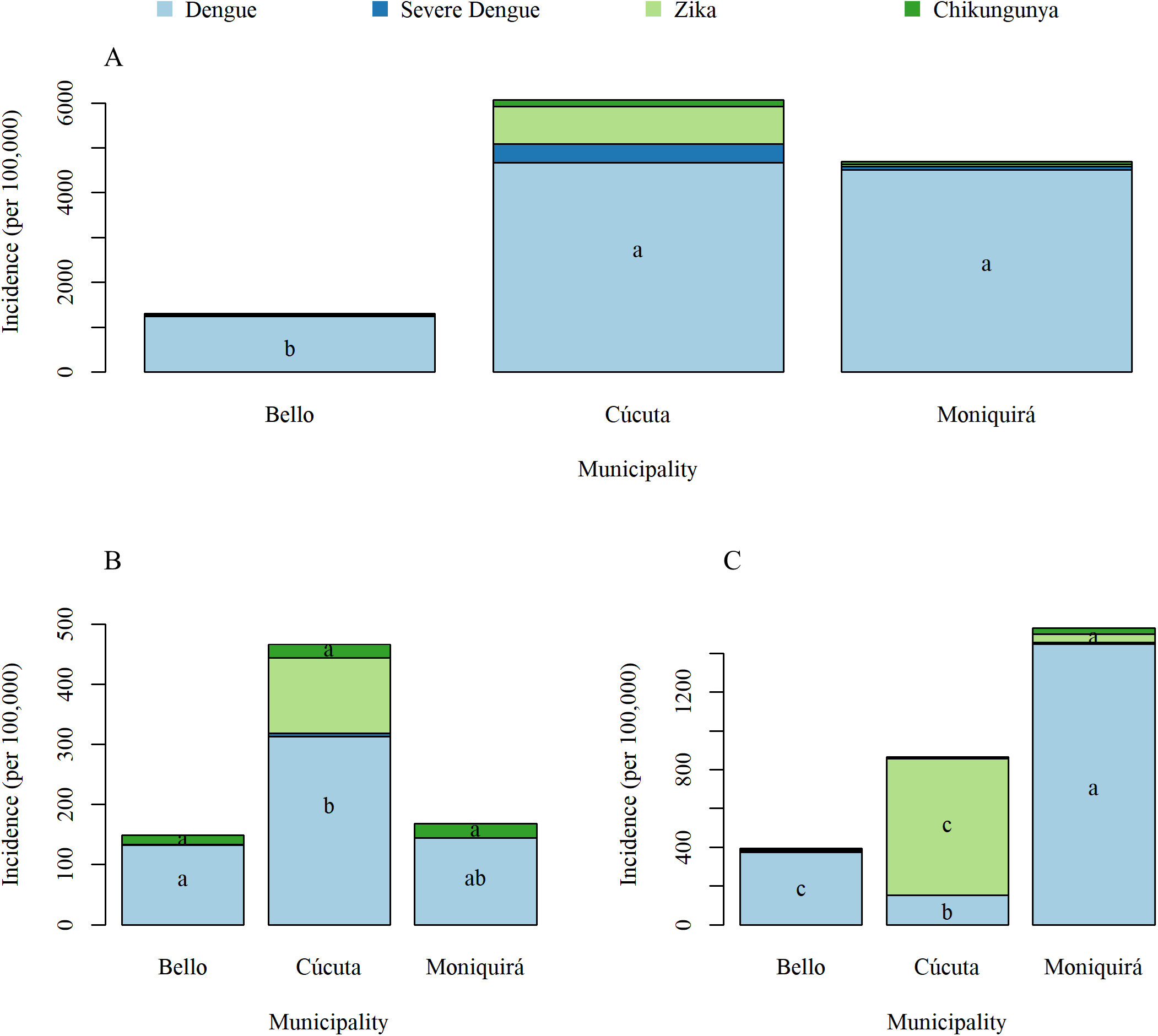
Total number of cases of dengue, severe dengue, chikungunya and Zika per 100,000 in the three municipalities. (A) 2007-2017, (B) 2015 and (C) 2016. The letters indicate significance of post-hoc Tukey test, where letters are different this indicates a significant difference (p =< 0.05).

The incidence data were also analysed separately for the years when the outbreaks of chikungunya and Zika were reported, 2015 and 2016, respectively (Fig 4B and 4C). This allowed for a more directly and meaningful comparison of the burden represented by these three diseases. Cases of chikungunya in 2015 were not significantly different between any of the municipalities (Fig 4B). However, Cúcuta had significantly higher incidence of Zika than both Bello (p= <0.001) and Moniquirá (p = <0.001) (Fig 4C). Interestingly, in the same year of the Zika outbreak (2016) each municipality had a significantly different number of dengue cases. Cúcuta had the lowest incidence of dengue, and Moniquirá the highest (Fig 4C). While Cúcuta had the highest incidence of dengue in previous years, in 2016 the same location experienced the lowest incidence of dengue accompanied by the highest incidence of Zika (Fig 4C).

### Patterns of disease incidence during 2007 to 2017

We compared the number of cases per 100,000 people in each year for dengue, severe dengue, chikungunya and Zika from 2007 to 2017 in Bello, Cúcuta and Moniquirá (Fig 5). The initial cases of Zika were first confirmed in Colombia in 2015, with cases reported in both Bello and Cúcuta from that year onwards. The first Zika case in Moniquirá was not confirmed until 2016. Whilst cases of Zika were relatively low in Bello and Moniquirá, Cúcuta experienced large outbreaks (Fig 5).

**Fig 5:**
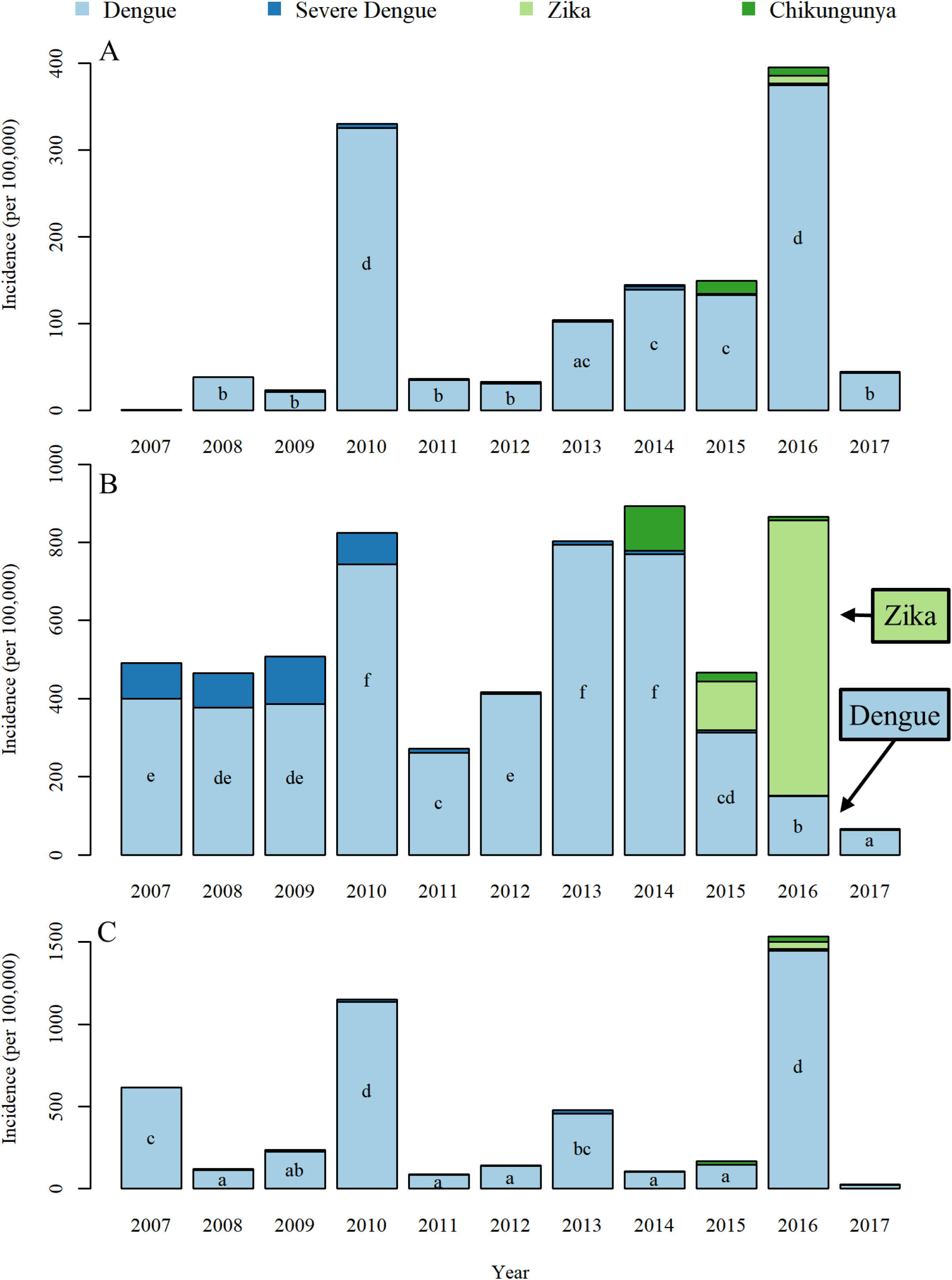
Annual cases of dengue, severe dengue, chikungunya and Zika per 100,000 people. (A) Bello, (B) Cúcuta and (C) Moniquirá. The letters indicate significance of post-hoc Tukey test, where letters are different this indicates a significant difference (p =< 0.05). Post-hoc Tukey tests show differences between years within each municipality. Note the spike in cases of dengue in 2010 in all three locations and the opposite trend in cases of Zika and dengue for Cúcuta (B) between 2015 and 2016.

The data analysed per location (Fig 4) and per year (Fig 5) suggested that Zika displaced dengue in Cúcuta from 2015 to 2016. The number of Zika cases per 100,000 population in 2015 were 125 and 705 in 2016 whilst dengue was present in 313 and 150 cases, respectively (Fig 5B). On the other hand, in Bello and Moniquirá, where the incidence of Zika was much lower (Bello; 1 in 2015 and 3 in 2016, Moniquirá; 0 in 2015 and 42 in 2016), there was an incremental trend for dengue during this same transition from 2015 to 2016 (Fig 5A and 5C). Following the significantly high dengue incidence in Bello and Moniquirá in 2016, the incidence stabilised, and the incidence reported in 2017 were not statistically different to that of years prior to 2016. This was not the case however in Cúcuta where incidence of dengue continued to fall, with the lowest incidence of the study period observed in 2017 (Fig 5B). In 2017 incidence of Zika was also much lower with only 3 cases per 100,000 people reported in Cúcuta in 2017 (Fig 5A and 5C).

### Effect of climate on disease incidence

The geographical settings for the three locations studied here Bello, Cúcuta and Moniquirá (Table 1) determine three different climate systems – ecosystems. These three different ecosystems are expected to establish contrasting behavioural patterns for the mosquito vector species that transmit dengue, Zika and chikungunya. The results presented in this section related to the quantitative climate variables as explained in Methods and inferred here to be proxy determinants of disease transmission by *Ae. aegypti*. We initially summarised the climate variables of each municipality from 2007-2017 as a breakdown of smooth moving averages (SMA) of 12-week periods (Fig 6). All parameters used here for temperature (Tmax, Tmin, Tavg) and wind speed (WSmax, WSmin, WSavg) were highest in Cúcuta. Bello had the lowest wind speed and highest relative humidity (Havg). There was a large fluctuation of 8°C in Tmax and 5°C in Tavg in 2011 in all three locations (approximately week 220, Fig 6). Otherwise the recorded climatic variables fluctuate within a similar range throughout these 11 years.

**Fig 6:**
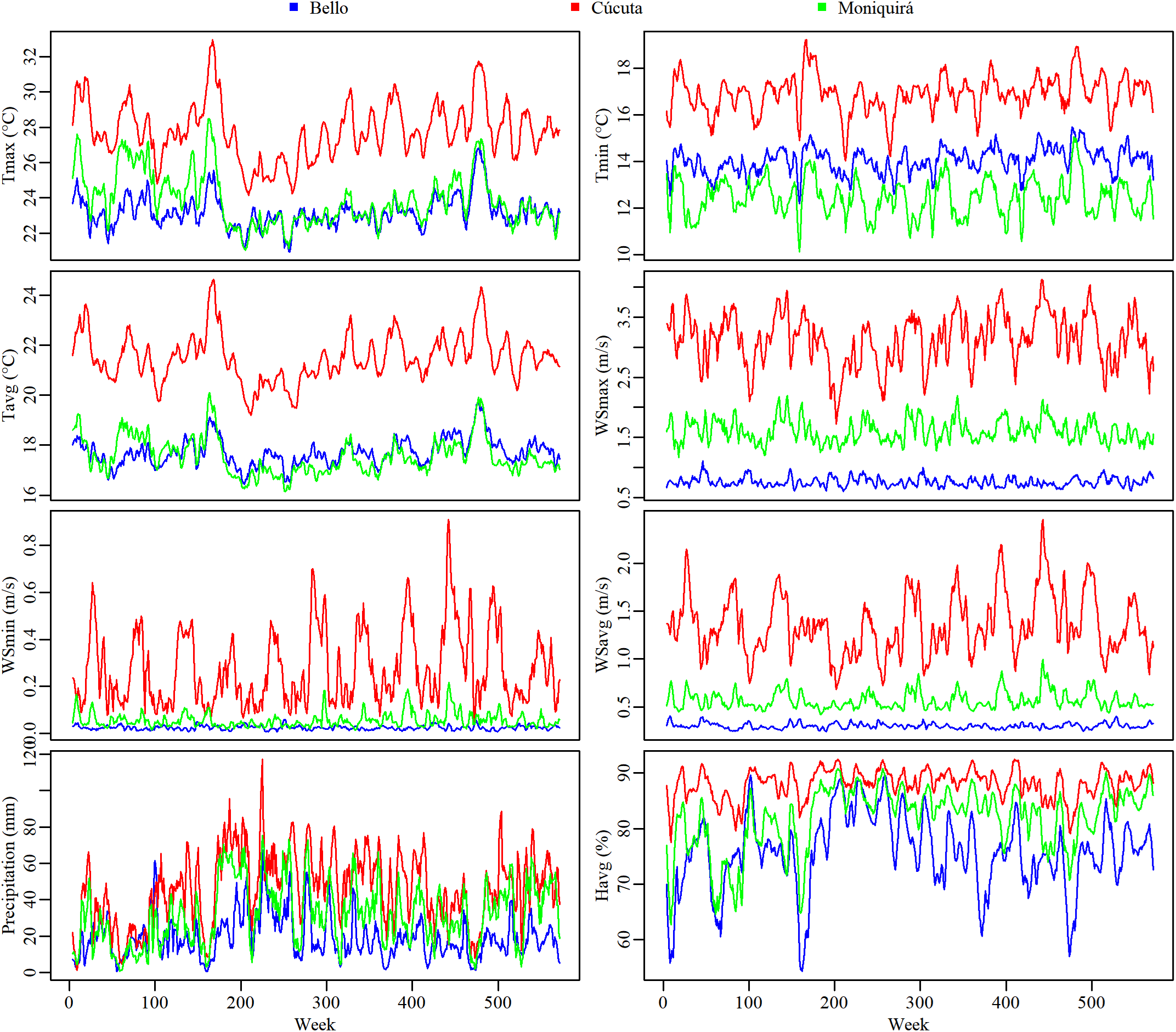
Time series data for climate in the municipalities of Bello, Cúcuta and Moniquirá between 2007 and 2017. Time series of maximum temperature (Tmax), minimum temperature (Tmin), average temperature (Tavg), maximum wind speed (WSmax), minimum wind speed (WSmin), average wind speed (WSavg), precipitation and average relative humidity (Havg), for Bello, Cúcuta and Moniquirá between 2007 and 2017.

We explored potential relationships between the time series climate data and total disease incidence in Bello, Cúcuta and Moniquirá at shorter 4 and 8-week time lags using a generalised additive model (GAM) as explained in Methods. The estimates from the quasipoisson GAM explained 57.6% of the variance in total disease incidence over time (Table 2). Effective degrees of freedom (*edf*) close to 1 represent relation close to linearity while high *edf* values for the smooth terms suggest that the relationship between climatic variables and disease incidence is non-linear. The GAM identified significant relationships between disease incidence and precipitation at 4 and 8-week lags, average humidity (4-week lag), minimum temperature (4 and 8-week lags), average temperature (8-week lag), maximum wind speed (4 and 8-week lag) and average wind speed (4-week lag) (Table 2).

**Table 2:**
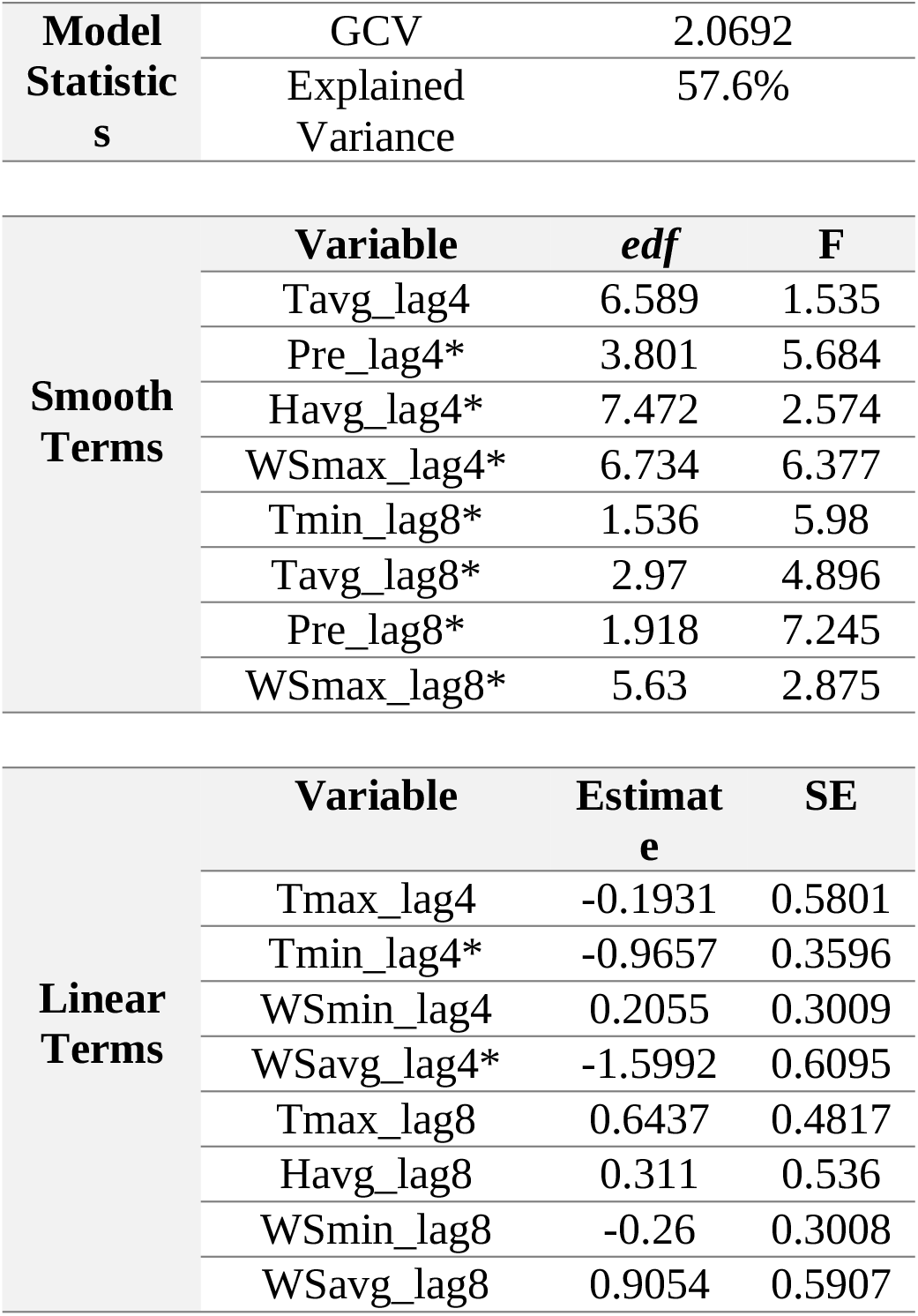
Quasi-GAM model estimates of the effects of climate variables on total disease incidence in Bello, Cúcuta and Moniquirá. Variable abbreviations as explained in Methods. GCV is the minimised generalised cross-validation. GCV was used for smoothness selection. F: F-statistic. SE: standard error of the mean. (*) Significant variable at the 0.001 level.

|  |  |  |
| --- | --- | --- |
| **Model Statistics** | GCV | 2.0692 |
|  | Explained Variance | 57.6% |

|  |  |  |  |
| --- | --- | --- | --- |
| **Smooth Terms** | **Variable** | **edf** | **F** |
|  | Tavg_lag4 | 6.589 | 1.535 |
|  | Pre_lag4* | 3.801 | 5.684 |
|  | Havg_lag4* | 7.472 | 2.574 |
|  | WSmax_lag4* | 6.734 | 6.377 |
|  | Tmin_lag8* | 1.536 | 5.98 |
|  | Tavg_lag8* | 2.97 | 4.896 |
|  | Pre_lag8* | 1.918 | 7.245 |
|  | WSmax_lag8* | 5.63 | 2.875 |
| **Linear Terms** | **Variable** | **Estimate** | **SE** |
|  | Tmax_lag4 | -0.1931 | 0.5801 |
|  | Tmin_lag4* | -0.9657 | 0.3596 |
|  | WSmin_lag4 | 0.2055 | 0.3009 |
|  | WSavg_lag4* | -1.5992 | 0.6095 |
|  | Tmax_lag8 | 0.6437 | 0.4817 |
|  | Havg_lag8 | 0.311 | 0.536 |
|  | WSmin_lag8 | -0.26 | 0.3008 |
|  | WSavg_lag8 | 0.9054 | 0.5907 |

The climate variables significantly correlated to total disease incidence as presented in Table 2 were further investigated by plotting the smoothed variance of the latter against the ranges covered for each climate variable (Fig 7). The analysis of the three most significant climatic contributors to total disease incidence -temperature, wind speed, and precipitation – delineated clear trends on how climate affected disease transmission. Increasing Tmin from 8°C to 16°C, at either 4 or 8-week, reduced the total disease incidence while the average temperatures (Tavg) up to 24°C contributed to incremental levels of disease (Fig 7A-C). This could be an indication of the need for a range of low minimal temperatures to adjust the average values at which disease transmission is more efficient.

**Fig 7:**
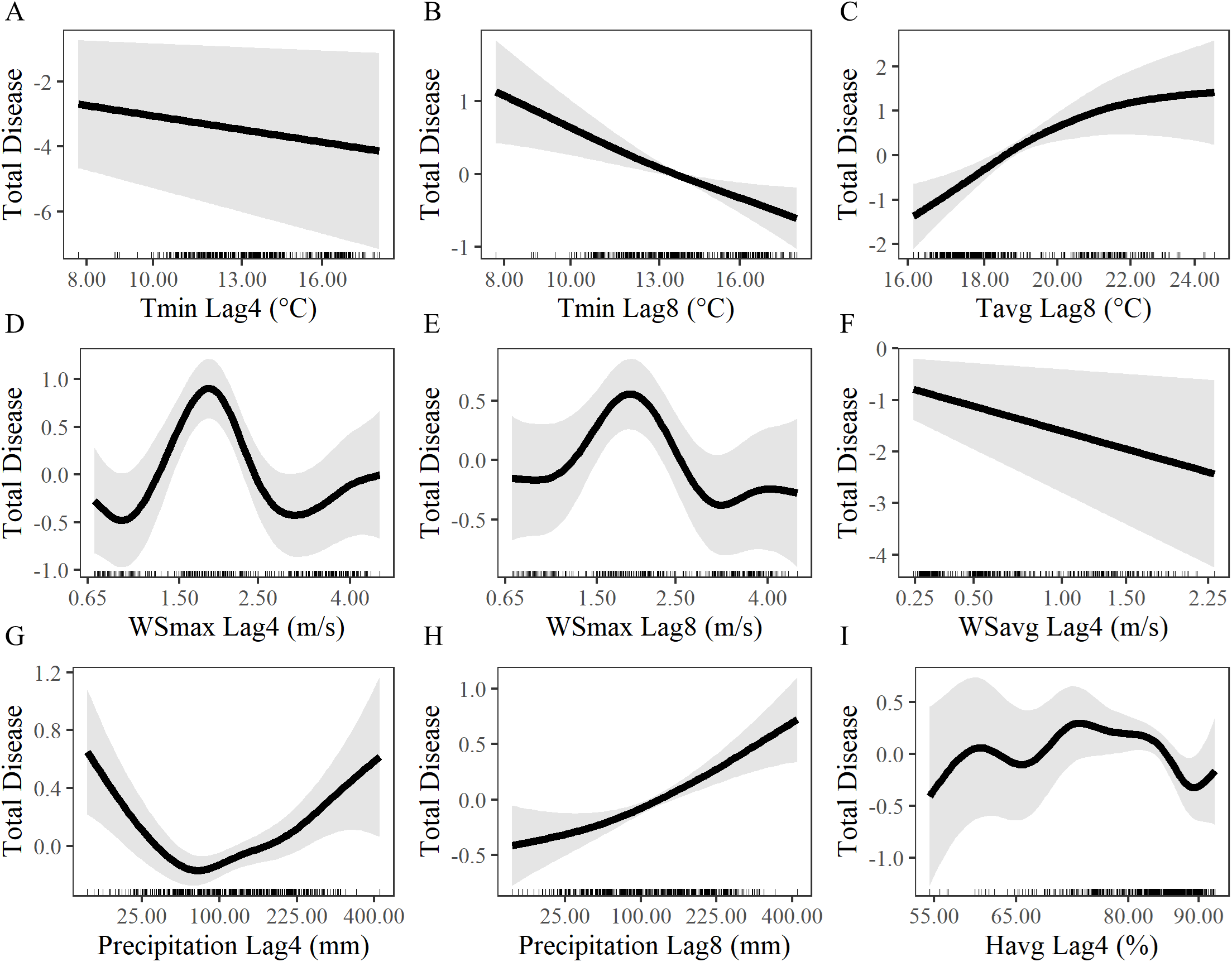
Climate determinants of total disease incidence. GAM estimated relationships between relative disease risk and minimum temperature (Tmin) with 4 (A) and 8-week (B) time lags, average temperature (Tavg) with 8-week time lag (C), maximum wind speed (WSmax) with 4 (D) and 8-week (E) time lags, precipitation with 4 (G) and 8-week (H) time lags and average humidity (Havg) with 4 week time lag (I).

Increasing wind speed above 1 m/s was associated by an increase in disease incidence with a peak around 2 m/s after which disease incidence declined until around 3 m/s where a small rise in disease risk can also be seen. This relationship between maximum wind speed and disease incidence was observed after both 4 and 8-week time lags (Fig 7D-E). The decisive influence on wind speed was substantiated by the negative effect on disease incidence at incremental average wind speeds (Fig 7F). Precipitation showed a positive relationship with disease incidence above 90 mm at 4-week time lags and above 25 mm for the data with 8-week time lag (Fig 7G-H). The high level of non-linearity shown for the relationship between average humidity and disease incidence (*edf* = 7.5) (Table 2) is detailed in Fig 7I. Disease incidence increased as humidity increased between 55-60%. This was followed by a slight decrease and plateau between 60-70%, a more rapid increase was observed between 70 and 75% above which disease incidence begins to decline (Fig 7I).

### Socio-economic profiles

We followed a holistic approach by further including socio-economic data for these three locations in the investigation for modifiers to the disease transmission of dengue, Zika and chikungunya. The overall multidimensional poverty index (see Methods) was lowest in Bello at 14.2. Cúcuta and Moniquirá had similar MPIs at 25.7 and 27.1 respectively (Table 3).

**Table 2:** **Socio-economic variables for the three municipalities of Bello, Cúcuta and Moniquirá overall and for each individual indicator**.

| Indicator | Total by Municipality (%) |  |  |
| --- | --- | --- | --- |
|  | Bello | Cúcuta | Moniquirá |
| **Socio-economic Status** |  |  |  |
| Multidimensional Poverty Measure <sup>(a)</sup> | 14.2 | 25.7 | 27.1 |
| **Education** |  |  |  |
| Illiteracy | 4.4 | 8.4 | 15.3 |
| Low Educational Achievement | 36.5 | 46.7 | 59.4 |
| **Childhood and Youth** |  |  |  |
| School lag/failure | 13.2 | 17 | 11.1 |
| School absence | 2.8 | 4.6 | 3.8 |
| Barriers to early childhood services | 1.8 | 2.2 | 1.6 |
| Child labour | 0.5 | 1.1 | 1.2 |
| **Health** |  |  |  |
| No health insurance | 19.2 | 17.5 | 12.4 |
| Barriers to health services | 2.8 | 5.2 | 2.9 |
| **Employment** |  |  |  |
| Informal work | 72.7 | 87 | 85.7 |
| Dependency rate | 25.4 | 34.3 | 29.2 |
| **Housing Conditions** |  |  |  |
| No access to improved water | 5.6 | 3.9 | 20.4 |
| Inadequate excreta disposal | 5.5 | 5.8 | 12.1 |
| Inappropriate flooring material | 0.7 | 3.6 | 4.3 |
| Inappropriate wall exterior | 1.4 | 6.1 | 0.6 |
| Critical overcrowding | 5.6 | 16.4 | 4.7 |

(a) Calculated using the data in S2 Table. Households with values of > 33.3% in any indicators are classed as poor.

The overall MPIs in Cúcuta and Moniquirá were similar. However, there were differences in specific poverty measures relevant to the transmission of vector borne viral diseases. Cúcuta had higher rates of overcrowding (16.4%), barriers to both childhood and youth services (2.2%) and healthcare services (5.2%) and inappropriate exterior wall material (6.1%) (Table 3). The findings also pointed to other socio-economic indexes that also affect general health and well-being in Moniquirá: inadequate excreta disposal (sanitation) (12.1%), no access to an improved water source (20.4%), illiteracy (15.3%) and low education achievement (59.4%) were all highest in Moniquirá (Table 3).

In order to introduce the socio-economic data into the analyses undertaken with the epidemiological and climatic data we carried out a dimensionality reduction and correlation with a principal component analysis (PCA). Importantly there was a clear separation of the three municipalities along both dimensions PC1 and PC2 that together integrates 89.9% of the compiled parameters (Fig 8). This approach made apparent a discriminatory set of factors both climatic and socio-economic for all three locations. Cúcuta had an extensive combination of climate factors (i.e. wind speed and temperature) that together with school absence, dependency, overcrowding, wall material and school failure are potential modifiers of dengue, Zika and chikungunya risk in this municipality. Interestingly, Moniquirá showed mainly socio-economic variables (i.e. water source, sanitation, illiteracy, low education, flooring material, child labour, high MPI, informal work) to be potential modifiers for disease risk. On the other hand, Bello had mainly climate variables as potential modifiers of disease transmission – average humidity, precipitation and elevation – with only health insurance as a socio-economic factor.

**Fig 8:**
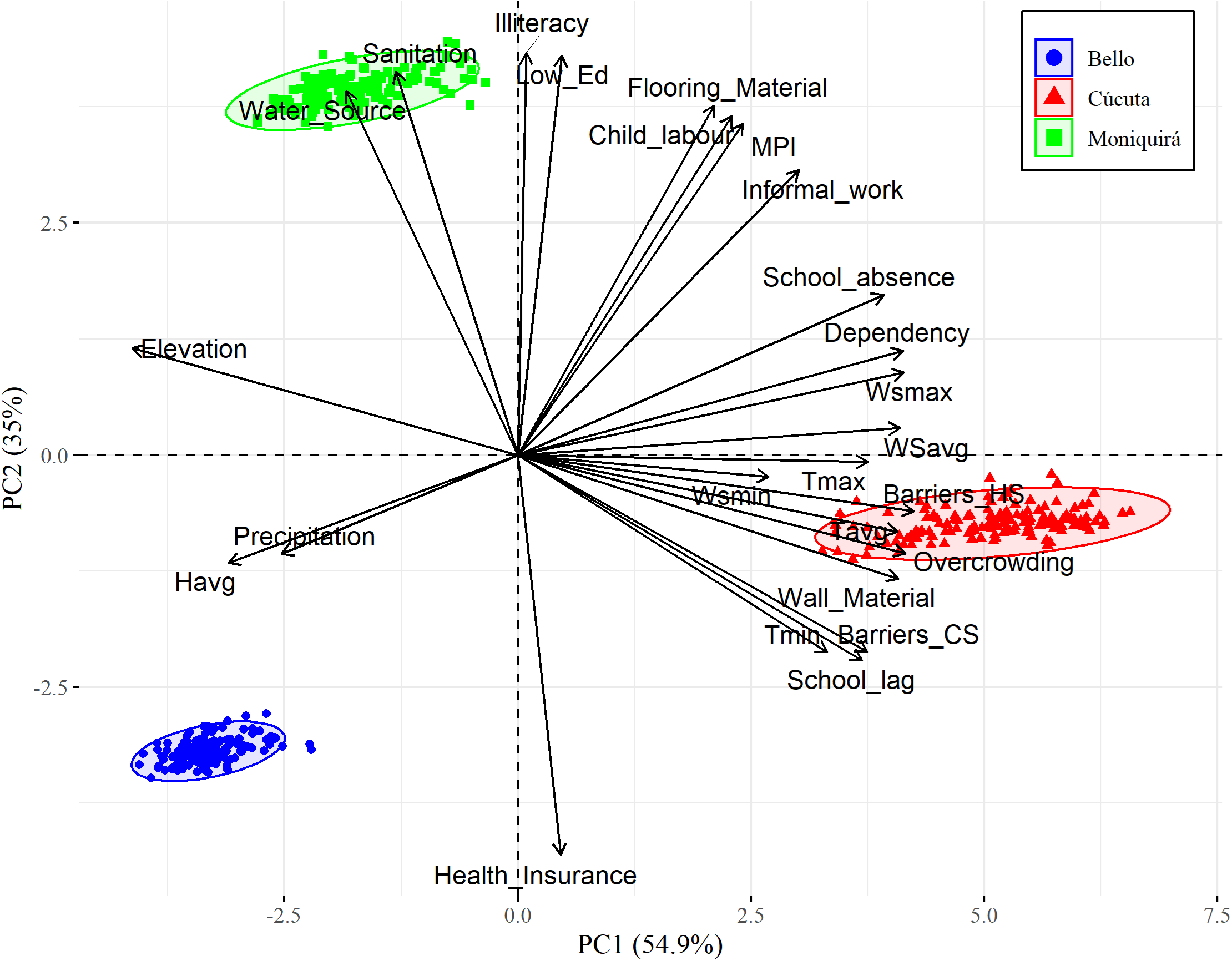
Principal component analysis for the both socio-economic and climate variables. Socio-economic variables: no access to improved water source (Water_Source), inadequate disposal for excreta (Sanitation), illiteracy, low educational achievement (Low_Ed), inappropriate flooring material (Flooring_Material), child labour, multidimensional poverty index (MPI), informal work, school absence, dependency rate (Dependency), barriers to health services (Barriers_HS), critical overcrowding, inappropriate wall material (Wall_Material), barriers to early childhood services (Bariers_CS), no health insurance (Health_Insurance). Climate variables: elevation, maximum wind speed (WSmax), average wind speed (WSavg), maximum temperature (Tmax), minimum wind speed (WSmin), average temperature (Tavg), precipitation (Pre) and average humidity (Havg). The length of the arrows represents the contribution of each variable.

## Discussion

This study set out to determine the longitudinal dynamics of three *Aedes* arboviral diseases co-circulating in three regions of Colombia over a 11-year period between 2007 and 2017. We found significant differences in the burden of viruses among the three municipalities studied. Bello had the lowest level of disease incidence across all diseases. Cúcuta had the highest incidence of severe dengue (2007-2017) and Zika (2016) and the highest overall disease incidence. In addition to climatic factors the burden of these vector borne diseases in Cúcuta can be compounded by local current socio-political dynamics. Cúcuta is on Colombia’s border with Venezuela, a country which has faced an economic, political and health crisis in recent years [42,43]. The humanitarian crisis has led to large migration of Venezuelan citizens and refugees to neighbouring countries, with Colombia receiving the highest number of Venezuelan migrants. The number of Venezuelan migrants in Colombia increased from 48,714 in 2015 to 600,000 in 2017 [44]. This has had a significant impact on public health in Colombia, with infectious and vector-borne diseases particularly effected [45–50].

The arrival of chikungunya and Zika in 2015 established a co-transmission of three different arboviruses by *Ae. aegypti*. Unexpectedly, a reduction in dengue cases was found in parallel to the spike in Zika cases in the year of the Zika epidemic of 2016 in Cúcuta. The same location in the years prior to the Zika outbreak had consistently presented high incidences of dengue. Moreover, in Cúcuta the incidence of both dengue and severe cases of dengue were lower in 2017, the year following the Zika epidemic, than in any of the 10 years prior. The decline of dengue following Zika reported in this study agrees with the overall decline in dengue incidence across the whole of Colombia [51,52] and has also been observed in other dengue endemic countries across the Americas [52,53]. In 2017 the total number of dengue cases across the Americas was lower than any of the 10 previous years [54] with a 73% decline between 2016 and 2017 alone [53].

Changes in epidemiological surveillance systems can cause the identification of inaccurate patterns of disease incidence. However, we did not observe any indication of significant changes to the surveillance system used to report *Aedes* borne viruses to SIVIGILIA, the database used here. However, the circulation of multiple viruses in the same localities at the same time does provide challenges for surveillance systems. Clinical presentation of Zika is very similar to that of dengue [22,55] and this can cause cases to be misidentified when laboratory testing is not conducted. We note that although the cases analysed in this study are all confirmed cases, confirmation is not always done by laboratory testing but also by epidemiological links. Misidentification could therefore explain the increase in dengue that was observed in Bello and Moniquirá in 2016, where low incidence of Zika was reported. Increased dengue incidence in 2015 and 2016 in other regions has also been reported and attributed to potential misidentification of Zika [53].

Coinfection of the primary vector *Ae. aegypti* with multiple arboviruses (i.e. DENV, CHIKV, ZIKV) has been reported following laboratory exposure [56–61]. *Aedes* mosquitoes have also been shown capable of transmitting more than one arbovirus in a single biting event [57,61,62]. Although coinfection has yet to be found in wild *Ae. aegypti* [62]. Coinfection of multiple *Aedes* borne viruses has been reported in mammalian hosts including humans [63– 69]. In Colombia patients have been diagnosed with DENV-CHIKV [48,70,71], DENV-ZIKV [48,71], CHIKV-ZIKV [48,71] and DENV-CHIKV-ZIKV [22,49] coinfections. However, the frequency of DENV-ZIKV co-infections seems low at 8.8% [71]. DENV and ZIKV co-transmission in mice through the bite of *Ae. aegypti* mosquitoes showed preferential transmission of ZIKV [61].

Host cross-immunity of ZIKV and DENV could have been a contributing factor in the dengue declines observed in this study. The observed decline in dengue cases following Zika outbreaks reported within this study and in others across the Americas suggest that there may be cross-immunity between ZIKV and DENV in humans [51–53,72–98]. Flavivirus immunity involves a T cell response and studies have reported cross-reactivity of CD4+ and CD8+ T cells to both DENV and ZIKV [99–103]. Cross-reactivity of antibodies and T cells and cross-immunity from Zika, although not necessarily conferring cross-protection, has been presented as the most probable reason explaining the decline of dengue across the Americas in 2017 [53].

Specific climatic factors associated here significantly affected disease incidence. This was particularly evident for Cúcuta, the municipality with the highest disease burden. We found significant co-relationship between average temperature and wind speed with disease transmission, with a peak at around 2m/s, consistent with findings of previous studies [104– 107]. *Ae. aegypti* has a small flight range of 200 m and high wind speeds reduce mosquito flight distances while low winds mean reduced dispersion of mosquitoes. We found the optimum wind speed to be around 2 m/s which is in line with current knowledge of mosquito flight [104,108,109]. Climate variables can be used to build predictive models to anticipate when outbreaks of dengue, chikungunya and Zika are likely to occur [110–113]. This is useful in the prioritisation of vector-control resources. Recent modelling studies have reported an increase in the incidence and geographical spread *Ae. aegypti* borne viruses when using climate change simulation models [114–118]. This highlights the importance of consideration of environmental factors when assessing risk of vector-borne disease [119].

We report differences in measurements of socio-economics between Bello, Cúcuta and Moniquirá. Bello, the municipality with the lowest burden of *Aedes* borne viruses also had the lowest poverty index, whilst Cúcuta and Moniquirá were much higher in both disease incidence and multidimensional poverty. Higher incidence of *Aedes* borne disease has been associated with lower socio-economic status and higher poverty levels [120–127]. Cúcuta had the highest rate of critical overcrowding. Overcrowding has been reported to be an important contributing factor to dengue incidence [119–121,128]. Inadequate sanitation, and no access to improved sources of water are both well-known contributing factors in increasing burden of *Aedes* borne disease due to the ecology of *Aedes* mosquitoes [26,119,123–125,129]. These socio-economic risk factors were highest in Moniquirá where there were also high levels of low educational achievement and illiteracy. Illiteracy and low educational level have previously been associated with increased vulnerability to dengue in Colombia and Brazil [130,131].

Heading into the COVID-19 2020 global pandemic, 2019 saw the highest number of dengue cases in The Americas for twenty years [132]. The factors that can exasperate the spread of COVID-19 (e.g. poor sanitation, lack of access to clean water, overcrowding) are also contributing factors to arboviral diseases. Disease surveillance and control programs are being significantly impacted by the pandemic, and the consequences of this will not be clear for some time but the pandemic also places an extra burden on already fragile health systems. Lockdown and social distancing measures will be detrimental for *Aedes* control which often involves community focused activities (e.g. door to door) led by community health workers and vector control experts. The World Mosquito Programme for example halted all control activities involving community interaction (e.g. releasing Wolbachia positive *Ae. aegypti*) for three months. Social distancing can be difficult or impossible in poorer areas with high population densities. It is therefore more imperative than ever that we more fully understand the dynamics of these important diseases which are likely to escalate over the coming years.

Having different ecosystems Bello, Cúcuta and Moniquirá presented a valuable opportunity to explore longitudinal arboviral disease incidence over 11 years that encompassed a Zika epidemic. Chikungunya was the only disease for which incidence did not significantly differ between the three municipalities. Cúcuta had the greatest disease incidence having the most favourable climatic factors and greater poverty index but as it borders with Venezuela mass movement of people is also suggested to be a contributing factor. Climatic factors associated with disease incidence were precipitation, average humidity, temperature and wind speed. Co-transmission of dengue and Zika during the epidemic led to a significant reduction of dengue cases in Cúcuta where dengue had previously been high. This significant finding warrants further investigation. Where the poverty index was low, as in Bello, so was the disease incidence. Socio-economic factors such as barriers to health and childhood services, inadequate sanitation, poor housing and poor water supply were implicated as drivers of disease transmission. *Aedes aegypti* and *Ae. albopictus* are increasing their geographical range and climate change is predicted to alter the distribution of these vectors and hence disease risk. Arboviral epidemiology is further complicated by humanitarian crises (e.g. Venezuela) and the COVID-19 pandemic which reinforces the urgency for understanding the dynamics of these global health problems. Context dependent and actionable understanding of the drivers for disease transmission that consider local dynamics, both climatic and socio-economic, should contribute to the design of more effective vector mosquito control programmes.

## Supporting information

Climatological and sociological data

Demographic data

Disease incidence data ten year period

## Data Availability

All data necessary to repeat or validated part or whole of the results presented in this manuscript are within the main body or in the supplementary files of the manuscript.

## Acknowledgments

We acknowledge Dr Ashley Lyons from Liverpool Hope University for advice on statistical techniques.

## Supporting information

**S1 Table. Disease, climate and socio-economic variables used in the analysis**.

**S2 Table. Indicators used for the calculation of MPI in Colombia including their respective weighting**. Calculations are completed at household level.

**S3 Table. Disease incidence data at department level**. Yearly incidence of dengue, severe dengue, chikungunya and Zika per 100,000 people in each Colombian department from 2007-2017.

## References

1. World Health Organization. A global brief on vector-borne diseases. World Heal Organ. 2014. Available from: http://apps.who.int/iris/bitstream/10665/111008/1/WHO_DCO_WHD_2014.1_eng.pdf

2. Liu LE, Dehning M, Phipps A, Swienton RE, Harris CA, Klein KR. Clinical Update on Dengue, Chikungunya, and Zika: What We Know at the Time of Article Submission. Disaster Med Public Health Prep. 2017;11: 290–299. doi:10.1017/dmp.2016.144

3. Kalayanarooj S. Clinical manifestations and management of dengue/DHF/DSS. Trop Med Health. 2011;39: 83–87. doi:10.2149/tmh.2011-S10

4. Song BH, Yun SI, Woolley M, Lee YM. Zika virus: History, epidemiology, transmission, and clinical presentation. J Neuroimmunol. 2017;308: 50–64. doi:10.1016/j.jneuroim.2017.03.001

5. Duffy MR, Chen TH, Hancock WT, Powers AM, Kool JL, Lanciotti RS, et al. Zika virus outbreak on Yap Island, Federated States of Micronesia. N Engl J Med. 2009;360: 2536–2543. doi:10.1056/NEJMoa0805715

6. De Barros Miranda-Filho D, Martelli CMT, De Alencar Ximenes RA, Araújo TVB, Rocha MAW, Ramos RCF, et al. Initial description of the presumed congenital Zika syndrome. Am J Public Health. 2016;106: 598–600. doi:10.2105/AJPH.2016.303115

7. Chan JFW, Choi GKY, Yip CCY, Cheng VCC, Yuen KY. Zika fever and congenital Zika syndrome: An unexpected emerging arboviral disease. J Infect. 2016;72: 507– 524. doi:10.1016/j.jinf.2016.02.011

8. Powers AM, Logue CH. Changing patterns of chikunya virus: Re-emergence of a zoonotic arbovirus. J Gen Virol. 2007;88: 2363–2377. doi:10.1099/vir.0.82858-0

9. Bhatt S, Gething PW, Brady OJ, Messina JP, Farlow AW, Moyes CL, et al. The global distribution and burden of dengue. Nature. 2013;496: 504–507. doi:10.1038/nature12060

10. World Health Organization (WHO). Dengue and severe dengue. In: WHO Factsheets [Internet]. 2020 [cited 31 Mar 2020]. Available from: http://www.who.int/mediacentre/factsheets/fs117/en/

11. Baud D, Gubler DJ, Schaub B, Lanteri MC, Musso D. An update on Zika virus infection. Lancet. 2017;390: 2099–2109. doi:10.1016/S0140-6736(17)31450-2

12. Ho ZJM, Hapuarachchi HC, Barkham T, Chow A, Ng LC, Lee JMV, et al. Outbreak of Zika virus infection in Singapore: an epidemiological, entomological, virological, and clinical analysis. Lancet Infect Dis. 2017;17: 813–821. doi:10.1016/S1473-3099(17)30249-9

13. Chu DT, Ngoc VTN, Tao Y. Zika virus infection in Vietnam: current epidemic, strain origin, spreading risk, and perspective. Eur J Clin Microbiol Infect Dis. 2017;36: 2041–2042. doi:10.1007/s10096-017-3030-8

14. Ruchusatsawat K, Wongjaroen P, Posanacharoen A, Rodriguez-Barraquer I, Sangkitporn S, Cummings DAT, et al. Long-term circulation of Zika virus in Thailand: an observational study. Lancet Infect Dis. 2019;19: 439–446. doi:10.1016/S1473-3099(18)30718-7

15. Lourenço J, Monteiro M, Tomás T, Monteiro Rodrigues J, Pybus O, Rodrigues Faria N. Epidemiology of the Zika Virus Outbreak in the Cabo Verde Islands, West Africa. PLoS Curr. 2018;10. doi:10.1371/currents.outbreaks.19433b1e4d007451c691f138e1e67e8c

16. Wahid B, Ali A, Rafique S, Idrees M. Global expansion of chikungunya virus: mapping the 64-year history. Int J Infect Dis. 2017;58: 69–76. doi:10.1016/j.ijid.2017.03.006

17. Powell JR, Gloria-Soria A, Kotsakiozi P. Recent history of Aedes aegypti: Vector genomics and epidemiology records. Bioscience. 2018;68: 854–860. doi:10.1093/biosci/biy119

18. Morin CW, Comrie AC, Ernst K. Climate and dengue transmission: Evidence and implications. Environ Health Perspect. 2013;121: 1264–1272. doi:10.1289/ehp.1306556

19. Dickens BL, Sun H, Jit M, Cook AR, Carrasco LR. Determining environmental and anthropogenic factors which explain the global distribution of aedes aegypti and Ae. Albopictus. BMJ Glob Heal. 2018;3. doi:10.1136/bmjgh-2018-000801

20. Kraemer MUGG, Sinka ME, Duda KA, Mylne AQNN, Shearer FM, Barker CM, et al. The global distribution of the arbovirus vectors Aedes aegypti and Ae. Albopictus. Elife. 2015;4: e08347. doi:10.7554/eLife.08347

21. Lippi CA, Stewart-Ibarra AM, Franklin BajañaLoor ME, Dueñas Zambrano JE, Espinoza Lopez NA, Blackburn JK, et al. Geographic shifts in Aedes aegypti habitat suitability in Ecuador using larval surveillance data and ecological niche modeling: Implications of climate change for public health vector control. PLoS Negl Trop Dis. 2019;13. doi:10.1371/journal.pntd.0007322

22. Rodriguez-Morales AJ, Villamil-Gómez WE, Franco-Paredes C. The arboviral burden of disease caused by co-circulation and co-infection of dengue, chikungunya and Zika in the Americas. Travel Med Infect Dis. 2016;14: 177–179. doi:10.1016/j.tmaid.2016.05.004

23. Portilla Cabrera CV, Selvaraj JJ. Geographic shifts in the bioclimatic suitability for Aedes aegypti under climate change scenarios in Colombia. Heliyon. 2020;6. doi:10.1016/j.heliyon.2019.e03101

24. Ruiz-López F, González-Mazo A, Vélez-Mira A, Gómez GF, Zuleta L, Uribe S, et al. Presencia de Aedes (stegomyia) aegypti (Linnaeus, 1762) y su infección natural con el virus del dengue en alturas no registradas para Colombia. Biomedica. 2016;36: 303– 308. doi:10.7705/biomedica.v36i2.3301

25. Pan American Health Organization (PAHO), World Health Organization (WHO). PLISA Health Information Platform for the Americas. In: Core indicators [Internet]. 2019 [cited 1 Apr 2020]. Available from: https://www.paho.org/data/index.php/en/

26. Spiegel JM, Bonet M, Ibarra AM, Pagliccia N, Ouellette V, Yassi A. Social and environmental determinants of Aedes aegypti infestation in Central Havana: Results of a case-control study nested in an integrated dengue surveillance programme in Cuba. Trop Med Int Heal. 2007;12: 503–510. doi:10.1111/j.1365-3156.2007.01818.x

27. Granada Y, Mejía-Jaramillo A, Strode C, Triana-Chavez O, Granada Y, Mejía-Jaramillo AM, et al. A Point Mutation V419L in the Sodium Channel Gene from Natural Populations of Aedes aegypti Is Involved in Resistance to λ-Cyhalothrin in Colombia. Insects. 2018;9: 23. doi:10.3390/insects9010023

28. Instituto Nacional de Salud. Portal Sivigila. [cited 30 May 2019]. Available from: http://portalsivigila.ins.gov.co/sivigila/documentos/Docs_1.php

29. NASA Langley Research Center (LaRC) POWER Project. POWER Data Access Viewer. [cited 17 Jan 2020]. Available from: https://power.larc.nasa.gov/data-access-viewer/

30. Departamento Administrativo Nacional de Estadística. Proyecciones de población. 2019 [cited 2 Oct 2019]. Available from: https://www.dane.gov.co/index.php/estadisticas-por-tema/demografia-y-poblacion/proyecciones-de-poblacion

31. Alkire S, Santos ME. Measuring Acute Poverty in the Developing World: Robustness and Scope of the Multidimensional Poverty Index. World Dev. 2014;59: 251–274. doi:10.1016/j.worlddev.2014.01.026

32. DANE. Medida de Pobreza Multidimensional Municipal de Fuente Censal Boletín Técnico. 2020. Available from: https://www.dane.gov.co/files/investigaciones/condiciones_vida/pobreza/2018/informacion-censal/bt-censal-pobreza-municipal-2018.pdf

33. Thomas R. Data Analysis with R Statistical Software A Guidebook for Scientists. 2nd ed. Eco-explore; 2017.

34. W. N. Venables and B. D. Ripley. Modern Applied Statistics with S. New York: Springer; 2002. Available from: http://www.stats.ox.ac.uk/pub/MASS4

35. Torsten H, Frank B, Westfall P. Simultaneous Inference in General Parametric Models. Biometrical J. 2008;50: 346-363.

36. R Core Team. R: A language and environment for statistical computing. Vienna, Austria: R Foundation for Statistical Computing; 2020. Available from: https://www.r-project.org/

37. Zuur AF, Ieno EN, Walker NJ, Saveliev AA, Smith GM. Mixed Effects Models and Extensions in Ecology with R. 1st ed. New York: Springer; 2009.

38. Wood SN, Pya N, Saefken B. Smoothing parameter and model selection for general smooth models (with discussion). J Am Stat Assoc; 2016. pp. 1548–1575.

39. Fasiolo M, Nedellec R, Goude Y, Wood Simon N. Scalable visualisation methods for modern Generalized Additive Models. 2018. Available from: https://arxiv.org/abs/1707.03307

40. Kassambara A, Mundt F. factoextra: Extract and Visualize the Results of Multivariate Data Analyses Title. 2020. Available from: https://cran.r-project.org/package=factoextra

41. Instituto de Hidrología M y EA. Características Climatológicas (1981 - 2010). 2014 [cited 2 Oct 2019]. Available from: https://web.archive.org/web/20160815030620/whttp://www.ideam.gov.co/web/tiempo-y-clima/clima

42. Nelson RM. Venezuela’s Economic Crisis: Issues for Congress. Congr Res Serv Libr Congr. 2018. Available from: https://pdfs.semanticscholar.org/c60d/ab688b21f960f6ada089e1c3bce87395fb9d.pdf

43. The Lancet. The collapse of the Venezuelan health system. Lancet. 2018;391: 1331. doi:10.1016/S0140-6736(16)00277-4

44. International Organisation for Migration (IOM). Migration Trends in The Americas. 2019. Available from: https://www.iom.int/external/migration-trends-americas-bolivarian-republic-venezuela

45. Grillet ME, Hernández-Villena J V., Llewellyn MS, Paniz-Mondolfi AE, Tami A, Vincenti-Gonzalez MF, et al. Venezuela’s humanitarian crisis, resurgence of vector-borne diseases, and implications for spillover in the region. Lancet Infect Dis. 2019;19: 149–161. doi:10.1016/S1473-3099(18)30757-6

46. Page KR, Doocy S, Reyna Ganteaume F, Castro JS, Spiegel P, Beyrer C. Venezuela’s public health crisis: a regional emergency. Lancet. 2019;393: 1254–1260. doi:10.1016/S0140-6736(19)30344-7

47. Doocy S, Page KR, de la Hoz F, Spiegel P, Beyrer C. Venezuelan Migration and the Border Health Crisis in Colombia and Brazil. J Migr Hum Secur. 2019;7: 79–91. doi:10.1177/2331502419860138

48. Carrillo-Hernández MY, Ruiz-Saenz J, Villamizar LJ, Gómez-Rangel SY, Martínez-Gutierrez M. Co-circulation and simultaneous co-infection of dengue, chikungunya, and zika viruses in patients with febrile syndrome at the Colombian-Venezuelan border. BMC Infect Dis. 2018;18: 61. doi:10.1186/s12879-018-2976-1

49. Tuite AR, Thomas-Bachli A, Acosta H, Bhatia D, Huber C, Petrasek K, et al. Infectious disease implications of large-scale migration of Venezuelan nationals. J Travel Med. 2018;25. doi:10.1093/jtm/tay077

50. Torres JR, Castro JS. Venezuela’s migration crisis: A growing health threat to the region requiring immediate attention. J Travel Med. 2019;26. doi:10.1093/jtm/tay141

51. Rico-Mendoza A, Porras-Ramírez A, Chang A, Encinales L, Lynch R. Co-circulation of dengue, chikungunya, and Zika viruses in Colombia from 2008 to 2018. Rev Panam Salud Pública. 2019;43: 49. doi:10.26633/rpsp.2019.49

52. Borchering RK, Huang AT, Mier-y-Teran-Romero L, Rojas DP, Rodriguez-Barraquer I, Katzelnick LC, et al. Impacts of Zika emergence in Latin America on endemic dengue transmission. Nat Commun. 2019;10: 1–9. doi:10.1038/s41467-019-13628-x

53. Perez F, Llau A, Gutierrez G, Bezerra H, Coelho G, Ault S, et al. The decline of dengue in the Americas in 2017: discussion of multiple hypotheses. Trop Med Int Heal. 2019;24: 442–453. doi:10.1111/tmi.13200

54. Pan American Health Organization (PAHO), World Health Organization (WHO). Epidemiological Alert: Dengue. 2018. Available from: http://www.paho.xn--orgpaho-qja6263e/WHO,2018

55. Beltrán-Silva SL, Chacón-Hernández SS, Moreno-Palacios E, Pereyra-Molina JÁ. Clinical and differential diagnosis: Dengue, chikungunya and Zika. Rev Médica del Hosp Gen México. 2018;81: 146–153. doi:10.1016/j.hgmx.2016.09.011

56. Nuckols JT, Huang Y-JJS, Higgs S, Miller AL, Pyles RB, Spratt HM, et al. Evaluation of simultaneous transmission of chikungunya virus and dengue virus type 2 in infected Aedes aegypti and Aedes albopictus (diptera: Culicidae). J Med Entomol. 2015;52: 447–451. doi:10.1093/jme/tjv017

57. Göertz GP, Vogels CBF, Geertsema C, Koenraadt CJM, Pijlman GP. Mosquito co-infection with Zika and chikungunya virus allows simultaneous transmission without affecting vector competence of Aedes aegypti. PLoS Negl Trop Dis. 2017;11. doi:10.1371/journal.pntd.0005654

58. Rückert C, Weger-Lucarelli J, Garcia-Luna SM, Young MC, Byas AD, Murrieta RA, et al. Impact of simultaneous exposure to arboviruses on infection and transmission by Aedes aegypti mosquitoes. Nat Commun. 2017;8: 1–9. doi:10.1038/ncomms15412

59. Le Coupanec A, Tchankouo-Nguetcheu S, Roux P, Khun H, Huerre M, Morales-Vargas R, et al. Co-infection of mosquitoes with chikungunya and dengue viruses reveals modulation of the replication of both viruses in midguts and salivary glands of Aedes aegypti mosquitoes. Int J Mol Sci. 2017;18: 1708. doi:10.3390/ijms18081708

60. Mourya DT, Gokhale MD, Majumdar TD, Yadav PD, Kumar V, Mavale MS. Experimental zika virus infection in aedes aegypti: Susceptibility, transmission & co-infection with dengue & chikungunya viruses. Indian J Med Res. 2018;147: 88–96. doi:10.4103/ijmr.IJMR_1142_17

61. Chaves BA, Orfano AS, Nogueira PM, Rodrigues NB, Campolina TB, Nacif-Pimenta R, et al. Coinfection with zika virus (ZIKV) and dengue virus results in preferential ZIKV transmission by vector bite to vertebrate host. J Infect Dis. 2018;218: 563–571. doi:10.1093/infdis/jiy196

62. Vogels CBF, Rückert C, Cavany SM, Perkins TA, Ebel GD, Grubaugh ND. Arbovirus coinfection and co-transmission: A neglected public health concern? PLoS Biol. 2019;17. doi:10.1371/journal.pbio.3000130

63. Iovine NM, Lednicky J, Cherabuddi K, Crooke H, White SK, Loeb JC, et al. Coinfection with zika and dengue-2 viruses in a traveler returning from Haiti, 2016: clinical presentation and genetic analysis. Clin Infect Dis. 2017;64: 72–75. doi:10.1093/cid/ciw667

64. Pessoa R, Patriota JV, De Lourdes De Souza M, Felix AC, Mamede N, Sanabani SS. Investigation into an outbreak of dengue-like illness in pernambuco, Brazil, revealed a cocirculation of Zika, Chikungunya, and dengue virus type 1. Medicine (Baltimore). 2016;95. doi:10.1097/MD.0000000000003201

65. Chang SF, Su CL, Shu PY, Yang CF, Liao TL, Cheng CH, et al. Concurrent isolation of chikungunya virus and dengue virus from a patient with coinfection resulting from a trip to Singapore. J Clin Microbiol. 2010;48: 4586–4589. doi:10.1128/JCM.01228-10

66. Waggoner JJ, Gresh L, Vargas MJ, Ballesteros G, Tellez Y, Soda KJ, et al. Viremia and Clinical Presentation in Nicaraguan Patients Infected With Zika Virus, Chikungunya Virus, and Dengue Virus. Clin Infect Dis. 2016;63: 1584–1590. doi:10.1093/cid/ciw589

67. Sardi SI, Somasekar S, Naccache SN, Bandeira AC, Tauro LB, Campos GS, et al. Coinfections of zika and chikungunya viruses in bahia, Brazil, identified by metagenomic next-generation sequencing. J Clin Microbiol. 2016;54: 2348–2353. doi:10.1128/JCM.00877-16

68. White SK, Mavian C, Elbadry MA, Beau De Rochars VM, Paisie T, Telisma T, et al. Detection and phylogenetic characterization of arbovirus dual-infections among persons during a chikungunya fever outbreak, Haiti 2014. PLoS Negl Trop Dis. 2018;12. doi:10.1371/journal.pntd.0006505

69. Chahar HS, Bharaj P, Dar L, Guleria R, Kabra SK, Broor S. Co-infections with chikungunya virus and dengue virus in Delhi, India. Emerg Infect Dis. 2009;15: 1077–1080. doi:10.3201/eid1507.080638

70. Mercado M, Acosta-Reyes J, Parra E, Pardo L, Rico A, Campo A, et al. Clinical and histopathological features of fatal cases with dengue and chikungunya virus co-infection in Colombia, 2014 to 2015. Eurosurveillance. 2016;21: 30244. doi:10.2807/1560-7917.ES.2016.21.22.30244

71. Mercado-Reyes M, Acosta-Reyes J, Navarro-Lechuga E, Corchuelo S, Rico A, Parra E, et al. Dengue, chikungunya and zika virus coinfection: Results of the national surveillance during the zika epidemic in colombia. Epidemiol Infect. 2019;147. doi:10.1017/S095026881800359X

72. Swanstrom JA, Plante JA, Plante KS, Young EF, McGowan E, Gallichotte EN, et al. Dengue virus envelope dimer epitope monoclonal antibodies isolated from dengue patients are protective against zika virus. MBio. 2016;7. doi:10.1128/mBio.01123-16

73. Collins MH, McGowan E, Jadi R, Young E, Lopez CA, Baric RS, et al. Lack of durable cross-neutralizing antibodies against zika virus from dengue virus infection. Emerg Infect Dis. 2017;23: 773–781. doi:10.3201/eid2305.161630

74. Zhang S, Kostyuchenko VA, Ng TS, Lim XN, Ooi JSG, Lambert S, et al. Neutralization mechanism of a highly potent antibody against Zika virus. Nat Commun. 2016;7: 1–7. doi:10.1038/ncomms13679

75. Langerak T, Mumtaz N, Tolk VI, Van Gorp ECM, Martina BE, Rockx B, et al. The possible role of cross-reactive dengue virus antibodies in Zika virus pathogenesis. PLoS Pathog. 2019;15. doi:10.1371/journal.ppat.1007640

76. Wen J, Shresta S. Antigenic cross-reactivity between Zika and dengue viruses: is it time to develop a universal vaccine? Curr Opin Immunol. 2019;59: 1–8. doi:10.1016/j.coi.2019.02.001

77. Zellweger RM, Prestwood TR, Shresta S. Enhanced Infection of Liver Sinusoidal Endothelial Cells in a Mouse Model of Antibody-Induced Severe Dengue Disease. Cell Host Microbe. 2010;7: 128–139. doi:10.1016/j.chom.2010.01.004

78. Balsitis SJ, Williams KL, Lachica R, Flores D, Kyle JL, Mehlhop E, et al. Lethal antibody enhancement of dengue disease in mice is prevented by Fc modification. PLoS Pathog. 2010;6. doi:10.1371/journal.ppat.1000790

79. Katzelnick LC, Gresh L, Halloran ME, Mercado JC, Kuan G, Gordon A, et al. Antibody-dependent enhancement of severe dengue disease in humans. Science. 2017;358: 929–932. doi:10.1126/science.aan6836

80. Salje H, Cummings DAT, Rodriguez-Barraquer I, Katzelnick LC, Lessler J, Klungthong C, et al. Reconstruction of antibody dynamics and infection histories to evaluate dengue risk. Nature. 2018;557: 719–723. doi:10.1038/s41586-018-0157-4

81. Screaton G, Mongkolsapaya J, Yacoub S, Roberts C. New insights into the immunopathology and control of dengue virus infection. Nat Rev Immunol. 2015;15: 745–759. doi:10.1038/nri3916

82. Dejnirattisai W, Jumnainsong A, Onsirisakul N, Fitton P, Vasanawathana S, Limpitikul W, et al. Cross-reacting antibodies enhance dengue virus infection in humans. Science. 2010;328: 745–748. doi:10.1126/science.1185181

83. Charles AS, Christofferson RC. Utility of a Dengue-Derived Monoclonal Antibody to Enhance Zika Infection In Vitro. PLoS Curr. 2016;8. doi:10.1371/currents.outbreaks.4ab8bc87c945eb41cd8a49e127082620

84. Bardina S V., Bunduc P, Tripathi S, Duehr J, Frere JJ, Brown JA, et al. Enhancement of Zika virus pathogenesis by preexisting antiflavivirus immunity. Science. 2017;356: 175–180. doi:10.1126/science.aal4365

85. Paul LM, Carlin ER, Jenkins MM, Tan AL, Barcellona CM, Nicholson CO, et al. Dengue virus antibodies enhance Zika virus infection. Clin Transl Immunol. 2016;5: 117. doi:10.1038/cti.2016.72

86. Castanha PMS, Nascimento EJM, Braga C, Cordeiro MT, De Carvalho O V, De Mendonça LR, et al. Dengue virus-specific antibodies enhance Brazilian Zika virus infection. J Infect Dis. 2017;215: 781–785. doi:10.1093/infdis/jiw638

87. Pantoja P, Pérez-Guzmán EX, Rodríguez I V., White LJ, González O, Serrano C, et al. Zika virus pathogenesis in rhesus macaques is unaffected by pre-existing immunity to dengue virus. Nat Commun. 2017;65: 1260–1265. doi:10.1038/ncomms15674

88. McCracken MK, Gromowski GD, Friberg HL, Lin X, Abbink P, De La Barrera R, et al. Impact of prior flavivirus immunity on Zika virus infection in rhesus macaques. PLoS Pathog. 2017;13. doi:10.1371/journal.ppat.1006487

89. Wang WH, Urbina AN, Wu CC, Lin CY, Thitithanyanont A, Assavalapsakul W, et al. An epidemiological survey of the current status of Zika and the immune interaction between dengue and Zika infection in Southern Taiwan. Int J Infect Dis. 2020;93: 151–159. doi:10.1016/j.ijid.2020.01.031

90. Kam YW, Lee CYP, Teo TH, Howland SW, Amrun SN, Lum FM, et al. Cross-reactive dengue human monoclonal antibody prevents severe pathologies and death from Zika virus infections. JCI insight. 2017;2. doi:10.1172/jci.insight.92428

91. Terzian ACB, Schanoski AS, De Oliveira Mota MT, Da Silva RA, Estofolete CF, Colombo TE, et al. Viral load and cytokine response profile does not support antibody-dependent enhancement in Dengue-Primed Zika Virus-infected patients. Clin Infect Dis. 2017;65: 1260–1265. doi:10.1093/cid/cix558

92. Ribeiro GS, Kikuti M, Tauro LB, Nascimento LCJ, Cardoso CW, Campos GS, et al. Does immunity after Zika virus infection cross-protect against dengue? Lancet Glob Heal. 2018;6: 140–141. doi:10.1016/S2214-109X(17)30496-5

93. Subramaniam KS, Lant S, Goodwin L, Grifoni A, Weiskopf D, Turtle L. Two Is Better Than One: Evidence for T-Cell Cross-Protection Between Dengue and Zika and Implications on Vaccine Design. Front Immunol. 2020;11: 517. doi:10.3389/fimmu.2020.00517

94. Pierson TC, Fremont DH, Kuhn RJ, Diamond MS. Structural Insights into the Mechanisms of Antibody-Mediated Neutralization of Flavivirus Infection: Implications for Vaccine Development. Cell Host Microbe. 2008;4: 229–238. doi:10.1016/j.chom.2008.08.004

95. Barba-Spaeth G, Dejnirattisai W, Rouvinski A, Vaney MC, Medits I, Sharma A, et al. Structural basis of potent Zika-dengue virus antibody cross-neutralization. Nature. 2016;536: 48–53. doi:10.1038/nature18938

96. Dejnirattisai W, Supasa P, Wongwiwat W, Rouvinski A, Barba-Spaeth G, Duangchinda T, et al. Dengue virus sero-cross-reactivity drives antibody-dependent enhancement of infection with zika virus. Nat Immunol. 2016;17: 1102–1108. doi:10.1038/ni.3515

97. Priyamvada L, Quicke KM, Hudson WH, Onlamoon N, Sewatanon J, Edupuganti S, et al. Human antibody responses after dengue virus infection are highly cross-reactive to Zika virus. Proc Natl Acad Sci U S A. 2016;113: 7852–7857. doi:10.1073/pnas.1607931113

98. Stettler K, Beltramello M, Espinosa DA, Graham V, Cassotta A, Bianchi S, et al. Specificity, cross-reactivity, and function of antibodies elicited by Zika virus infection. Science. 2016;353: 823–826. doi:10.1126/science.aaf8505

99. Grifoni A, Pham J, Sidney J, O’Rourke PH, Paul S, Peters B, et al. Prior Dengue Virus Exposure Shapes T Cell Immunity to Zika Virus in Humans. J Virol. 2017;91. doi:10.1128/jvi.01469-17

100. Wen J, Elong Ngono A, Angel Regla-Nava J, Kim K, Gorman MJ, Diamond MS, et al. Dengue virus-reactive CD8+ T cells mediate cross-protection against subsequent Zika virus challenge. Nat Commun. 2017;8: 1–11. doi:10.1038/s41467-017-01669-z

101. Wen J, Tang WW, Sheets N, Ellison J, Sette A, Kim K, et al. Identification of Zika virus epitopes reveals immunodominant and protective roles for dengue virus cross-reactive CD8+ T cells. Nat Microbiol. 2017;2: 17036. doi:10.1038/nmicrobiol.2017.36

102. Herrera BB, Tsai W-Y, Chang CA, Hamel DJ, Wang W-K, Lu Y, et al. Sustained Specific and Cross-Reactive T Cell Responses to Zika and Dengue Virus NS3 in West Africa. J Virol. 2018;92. doi:10.1128/jvi.01992-17

103. Lim MQ, Kumaran EAP, Tan HC, Lye DC, Leo YS, Ooi EE, et al. Cross-reactivity and anti-viral function of dengue capsid and NS3-specific memory t cells toward Zika Virus. Front Immunol. 2018;9: 2225. doi:10.3389/fimmu.2018.02225

104. Cheong YL, Burkart K, Leitão PJ, Lakes T. Assessing weather effects on dengue disease in Malaysia. Int J Environ Res Public Health. 2013;10: 6319–6334. doi:10.3390/ijerph10126319

105. Johansson MA, Cummings DAT, Glass GE. Multiyear climate variability and dengue -El Niño southern oscillation, weather, and dengue incidence in Puerto Rico, Mexico, and Thailand: A longitudinal data analysis. PLoS Med. 2009;6. doi:10.1371/journal.pmed.1000168

106. Peña-García VH, Triana-Chávez O, Arboleda-Sánchez S. Estimating Effects of Temperature on Dengue Transmission in Colombian Cities. Ann Glob Heal. 2017;83: 509–518. doi:10.1016/j.aogh.2017.10.011

107. Mordecai EA, Cohen JM, Evans M V., Gudapati P, Johnson LR, Lippi CA, et al. Detecting the impact of temperature on transmission of Zika, dengue, and chikungunya using mechanistic models. PLoS Negl Trop Dis. 2017;11. doi:10.1371/journal.pntd.0005568

108. Paskewitz SM. The Biology of Mosquitoes. Volume 1. Development, Nutrition and Reproduction. Am J Trop Med Hyg. 1995;52: 579–579. doi:10.4269/ajtmh.1995.52.579

109. Lu L, Lin H, Tian L, Yang W, Sun J, Liu Q. Time series analysis of dengue fever and weather in Guangzhou, China. BMC Public Health. 2009;9: 395. doi:10.1186/1471-2458-9-395

110. Descloux E, Mangeas M, Menkes CE, Lengaigne M, Leroy A, Tehei T, et al. Climate-based models for understanding and forecasting dengue epidemics. PLoS Negl Trop Dis. 2012;6. doi:10.1371/journal.pntd.0001470

111. Lowe R, Bailey TC, Stephenson DB, Graham RJ, Coelho CAS, Sá Carvalho M, et al. Spatio-temporal modelling of climate-sensitive disease risk: Towards an early warning system for dengue in Brazil. Comput Geosci. 2011;37: 371–381. doi:10.1016/j.cageo.2010.01.008

112. Zhu B, Wang L, Wang H, Cao Z, Zha L, Li Z, et al. Prediction model for dengue fever based on interactive effects between multiple meteorological factors in Guangdong, China (2008–2016). PLoS One. 2019;14. doi:10.1371/journal.pone.0225811

113. Jayaraj VJ, Avoi R, Gopalakrishnan N, Raja DB, Umasa Y. Developing a dengue prediction model based on climate in Tawau, Malaysia. Acta Trop. 2019;197: 105055. doi:10.1016/j.actatropica.2019.105055

114. Colón-González FJ, Fezzi C, Lake IR, Hunter PR. The Effects of Weather and Climate Change on Dengue. PLoS Negl Trop Dis. 2013;7: 2503. doi:10.1371/journal.pntd.0002503

115. Messina JP, Brady OJ, Pigott DM, Golding N, Kraemer MUG, Scott TW, et al. The many projected futures of dengue. Nat Rev Microbiol. 2015;13: 230–239. doi:10.1038/nrmicro3430

116. Xu Z, Bambrick H, Frentiu FD, Devine G, Yakob L, Williams G, et al. Projecting the future of dengue under climate change scenarios: Progress, uncertainties and research needs. PLoS Negl Trop Dis. 2020;14. doi:10.1371/journal.pntd.0008118

117. Messina JP, Brady OJ, Golding N, Kraemer MUG, Wint GRW, Ray SE, et al. The current and future global distribution and population at risk of dengue. Nat Microbiol. 2019;4: 1508–1515. doi:10.1038/s41564-019-0476-8

118. Lee H, Kim JE, Lee S, Lee CH. Potential effects of climate change on dengue transmission dynamics in Korea. PLoS One. 2018;13. doi:10.1371/journal.pone.0199205

119. Delmelle E, Hagenlocher M, Kienberger S, Casas I. A spatial model of socioeconomic and environmental determinants of dengue fever in Cali, Colombia. Acta Trop. 2016;164: 169–176. doi:10.1016/j.actatropica.2016.08.028

120. Zellweger RM, Cano J, Mangeas M, Taglioni F, Mercier A, Despinoy M, et al. Socioeconomic and environmental determinants of dengue transmission in an urban setting: An ecological study in Nouméa, New Caledonia. PLoS Negl Trop Dis. 2017;11. doi:10.1371/journal.pntd.0005471

121. Braga C, Luna CF, Martelli CMT, Souza WV de, Cordeiro MT, Alexander N, et al. Seroprevalence and risk factors for dengue infection in socio-economically distinct areas of Recife, Brazil. Acta Trop. 2010;113: 234–240. doi:10.1016/j.actatropica.2009.10.021

122. Johansson MA, Dominici F, Glass GE. Local and global effects of climate on dengue transmission in Puerto Rico. PLoS Negl Trop Dis. 2009;3: 382. doi:10.1371/journal.pntd.0000382

123. Costa JV, Donalisio MR, Silveira LV de A. Spatial distribution of dengue incidence and socio-environmental conditions in Campinas, São Paulo State, Brazil, 2007. Cad Saude Publica. 2013;29: 1522–1532. doi:10.1590/0102-311X00110912

124. Mocelin HJS, Catão RC, Freitas PSS, Prado TN, Bertolde AI, Castro MC, et al. Analysis of the spatial distribution of cases of Zika virus infection and congenital Zika virus syndrome in a state in the southeastern region of Brazil: Sociodemographic factors and implications for public health. Int J Gynecol Obstet. 2020;148: 61–69. doi:10.1002/ijgo.13049

125. Ali S, Gugliemini O, Harber S, Harrison A, Houle L, Ivory J, et al. Environmental and Social Change Drive the Explosive Emergence of Zika Virus in the Americas. PLoS Negl Trop Dis. 2017;11. doi:10.1371/journal.pntd.0005135

126. Raude J, Setbon M. The role of environmental and individual factors in the social epidemiology of chikungunya disease on Mayotte Island. Heal Place. 2009;15: 689– 699. doi:10.1016/j.healthplace.2008.10.009

127. Koyadun S, Butraporn P, Kittayapong P. Ecologic and sociodemographic risk determinants for dengue transmission in urban areas in Thailand. Interdiscip Perspect Infect Dis. 2012;2012. doi:10.1155/2012/907494

128. Soghaier MA, Himatt S, Osman KE, Okoued SI, Seidahmed OE, Beatty ME, et al. Cross-sectional community-based study of the socio-demographic factors associated with the prevalence of dengue in the eastern part of Sudan in 2011 Infectious Disease epidemiology. BMC Public Health. 2015;15: 1–6. doi:10.1186/s12889-015-1913-0

129. Carlton EJ, Liang S, McDowell JZ, Li H, Luo W, Remais J V. Regional disparities in the burden of disease attributable to unsafe water and poor sanitation in China. Bull World Health Organ. 2012;90: 578–587. doi:10.2471/BLT.11.098343

130. Hagenlocher M, Delmelle E, Casas I, Kienberger S. Assessing socioeconomic vulnerability to dengue fever in Cali, Colombia: Statistical vs expert-based modeling. Int J Health Geogr. 2013;12: 36. doi:10.1186/1476-072X-12-36

131. Siqueira-Junior JB, Maciel IJ, Barcellos C, Souza W V., Carvalho MS, Nascimento NE, et al. Spatial point analysis based on dengue surveys at household level in central Brazil. BMC Public Health. 2008;8: 361. doi:10.1186/1471-2458-8-361

132. Pan American Health Organization, World Health Organization. Epidemiological Update: Dengue. 2020. Available from: https://www.paho.org/en/documents/epidemiological-update-dengue-7-february-2020

